# LUCID: Lipid-featured Urine Cytology with Intelligent Detection for Non-Invasive Early Diagnosis of Bladder Cancer

**DOI:** 10.64898/2026.10.14.26365504

**Authors:** Zhe Hao, Jianlin Liu, Tinghe Fang, Yaoyao Zheng, Hao Hu, Shuai Hu, Xinyan Zhang, Liqun Zhou, Xun Chen, Lin Yao, Shuhua Yue

**Author notes:** These authors contributed equally (Zhe Hao, Jianlin Liu, Tinghe Fang, Yaoyao Zheng).

## Abstract

Leveraging its non-invasive and convenient nature, urinary cytology offers a way for the early diagnosis and surveillance of bladder cancer (BCa). However, its sensitivity remains substantially inferior to the histopathological gold standard, rendering it inadequate to support precise clinical diagnosis. Here, we developed Lipid-featured Urine Cytology with Intelligent Detection (LUCID), an automated platform integrating single-cell stimulated Raman scattering (SRS) imaging and explainable AI model for accurate BCa detection. Following a newly developed standardized urine-preprocessing protocol, 19,678 urinary exfoliated cells from 26 BCa patients and 54 non-BCa participants were imaged using two-color SRS microscopy to simultaneously visualize lipid and protein distributions. For cell-level analysis, a customized deep-supervised Cellpose segmentation algorithm and a YOLO-based detection model were developed to enable precise single-cell segmentation and intracellular lipid droplet (LD) identification, respectively. Totally 20 quantitative morphological and compositional features per cell were extracted and 10 related principal components were obtained via principal component analysis. These features were subsequently integrated into a multiple-instance learning (MIL)-XGBoosting model, which achieved robust diagnostic performance, with 92.31% sensitivity, 87.04% specificity, and an AUC of 0.940. We found that morphological and compositional features provided complementary diagnostic information, while LD-associated features provided primary discriminative information, supporting intracellular LD phenotypes as potential urinary cellular markers of BCa. More importantly, LUCID demonstrated exceptional accuracy in detecting early-stage and small tumors, and showed higher concordance than FISH in a preliminary paired subset (5/6 vs. 2/6). Together, LUCID establishes a chemical imaging–AI paradigm that transforms molecular urine cytology.

## Introduction

Bladder cancer (BCa) is among the most common malignancies of the urinary tract and the eighth most frequently diagnosed cancer worldwide, with an estimated 635,000 new cases and 228,000 deaths in 2024^1^. Approximately 80% of cases are non-muscle-invasive BCa (NMIBC; earlystages Ta, T1, and Tis) which have high recurrence rate and risk of progression to muscle-invasive BCa (MIBC; stages T2–4)^2^. The 5-year survival rate exceeds 85% for NMIBC^3^, decreases to approximately 50% for non-metastatic MIBC^4^, and is only approximately 10% in patients with visceral metastatic urothelial cancer^5^. Therefore, early detection and regular surveillance of BCa are critical. Current gold standard of diagnosis is histopathology of cystoscopic tissue biopsy, which is invasive, operator-dependent, and carries risks of infection. This challenge highlights the urgent need for non-invasive and accurate diagnostic alternatives.

In terms of non-invasive diagnostic methods, urine cytology is commonly used with high specificity but suffers from limited sensitivity (∼40%)^6,7^, likely due to subjective interpretation of the morphology of urinary exfoliated cells (UECs) without tissue architecture. To improve the sensitivity of BCa detection with UECs, assays based on specific protein markers, chromosomal aberrations, and DNA methylation have been developed. For instance, detection of NMP22, a BCa protein marker, had a specificity about 89% but a low sensitivity of about 57%^8^. The multi-target UroVysion fluorescence in situ hybridization (FISH) assay showed a pooled sensitivity of 65% and specificity of 81% in a recent meta-analysis^9^; sensitivity has been reported to decrease to approximately 41% for low-grade tumors^10^. The mass spectrometry-based DNA methylation test achieved 90% sensitivity and 83% specificity, but it suffers from complex testing and high costs^11^. The latest BLOOM platform, utilizing urinary hyaluronidase as a biomarker, achieves a sensitivity of 88.3% and an accuracy of 90%. It demonstrates excellent resistance to urine heterogeneity and is well-suited for bedside testing^12^. However, the current assay requires controlled incubation at 37 °C for optimal hyaluronidase activity, with ambient-temperature operation remaining a goal for further development. The intricate heterogeneity of gene and protein expression in BCa cells makes current diagnostic methods fall short of clinical requirements^13,14^. Therefore, it is imperative to develop a universally applicable method that can non-invasively detect BCa with both high specificity and sensitivity.

In addition to alterations in genes and proteins, dysregulated lipid metabolism has been recognized as a hallmark of human cancers^15-17^. Lipid droplets (LDs), as dynamic organelles involved in lipid storage and metabolic homeostasis, are increasingly recognized as important regulators of cancer-cell survival, stress adaptation, and tumor progression^18^. Specifically, both de novo synthesis and uptake of lipids are significantly upregulated in BCa^19,20^. Activation of the AKT/mTOR/SREBP-1c/FASN signaling axis in BCa promotes lipogenesis and is associated with enhanced LD accumulation^21,22^, whereas FABP5-mediated lipid metabolism contributes to BCa progression^23^. Thus, it is worth exploring whether the lipid features, for instance intracellular LDs, could be sensitive markers in UECs for BCa detection.

For intracellular lipid analysis, the commonly used fluorescence imaging relies on exogenous lipophilic dyes, which tend to bind to impurities present in urine, leading to non-specific staining. In addition, mass spectrometry imaging (MSI) is currently well-suited for histochemical imaging, but its limited spatial resolution restricts the sensitive detection of chemical information at the subcellular level. Notably, stimulated Raman scattering (SRS) microscopy has emerged as a powerful tool for subcellular chemical imaging, which is capable to map molecular chemical fingerprints by probing intrinsic bond vibrations without any labeling^24-26^. Specifically, by targeting the distinct C–H stretching vibrational signatures of lipid molecules, SRS enables label-free and quantitative profiling of lipid-rich structures, especially LDs, at subcellular spatial resolution. Over the past decade, SRS microscopy has been extensively utilized to investigate lipid metabolic reprogramming across diverse cancer types, yielding fundamental insights into the role of LDs in tumor progression^27-30^.

Furthermore, powered by deep learning algorithms, SRS has revolutionized rapid intraoperative tumor diagnosis, enabling highly accurate tissue assessment for brain and other solid tumors^31-33^. Building upon these tissue-level advancements, our previous work established SRS-based molecular cytology, which enables accurate diagnosis by leveraging single-cell biochemical composition in the absence of tissue architecture. Using this strategy, we detected metastatic gastric cancer cells in peritoneal lavage fluid^34^ and subsequently applied it to intraoperative margin assessment in pancreatic ductal adenocarcinoma^35^. However, adapting this SRS-based molecular cytology for the early detection of BCa via UECs introduces unprecedented challenges. First, compared to peritoneal lavage fluids and pancreatic margin brushing samples, human urine is a vastly more complex and heterogeneous matrix, with interfering impurities such as red blood cells, urinary crystals, protein flocculates, and bacteria, all of which hinder cell identification and contaminate the chemical vibrational signatures of target cells. Second, urinary exfoliated cells exhibit exceptionally high morphological heterogeneity, posing significant challenges for single-cell segmentation. Third, accurately extracting highly interpretable features, particularly those related to LDs from morphologically heterogeneous UECs remains challenging. Fourth, the proportion of malignant cells is markedly lower and heavily obscured by a predominant background of benign cells (e.g., squamous epithelial cells and leukocytes), making their identification extremely challenging.

In this study, we developed Lipid-featured Urine Cytology with Intelligent Detection (LUCID), a label-free molecular cytology framework for non-invasive BCa detection (**Fig. 1**). As illustrated in **Fig. 1a**, urine samples were subjected to standardized preprocessing to remove major interfering components and enrich UECs, followed by two-color SRS imaging to characterize their lipid- and protein-related chemical information. Subsequently, Deep Supervision (DS)-Cellpose and YOLOv5 were used for single-cell segmentation and LD detection (**Fig. 1b**), respectively, enabling the quantitative extraction and selection of single-cell morphological and compositional features. Finally, the extracted features were transformed by principal component analysis (PCA) and analyzed using a multiple-instance learning (MIL)-XGBoosting framework to identify BCa-associated significant marker cells (**Fig. 1c**), whose predicted probabilities were aggregated by max pooling to generate patient-level BCa predictions. A panel of feature combinations, including morphology, Lipid & Protein, LD, and their comprehensive integrations (e.g., morphology & LD), was independently trained and compared within the proposed framework. With leave-one-out cross-validation, our LUCID method (based on morphology & composition features) ultimately achieved 92.31% sensitivity, 87.04% specificity, and an area under the curve (AUC) of 0.940 for overall BCa (n=80), outperforming all other feature combinations tested. Feature-ablation analysis further identified LD-associated features as a major source of discriminative information, while their integration with broader compositional and morphological features jointly enhanced the sensitivity and specificity of BCa diagnosis. More importantly, LUCID also demonstrated exceptional diagnostic accuracy for early-stage BCa, that is, 90.91% concordance with pathological diagnoses. In summary, LUCID may offer new opportunities for non-invasive and accurate early diagnosis of BCa.

**Fig. 1.**
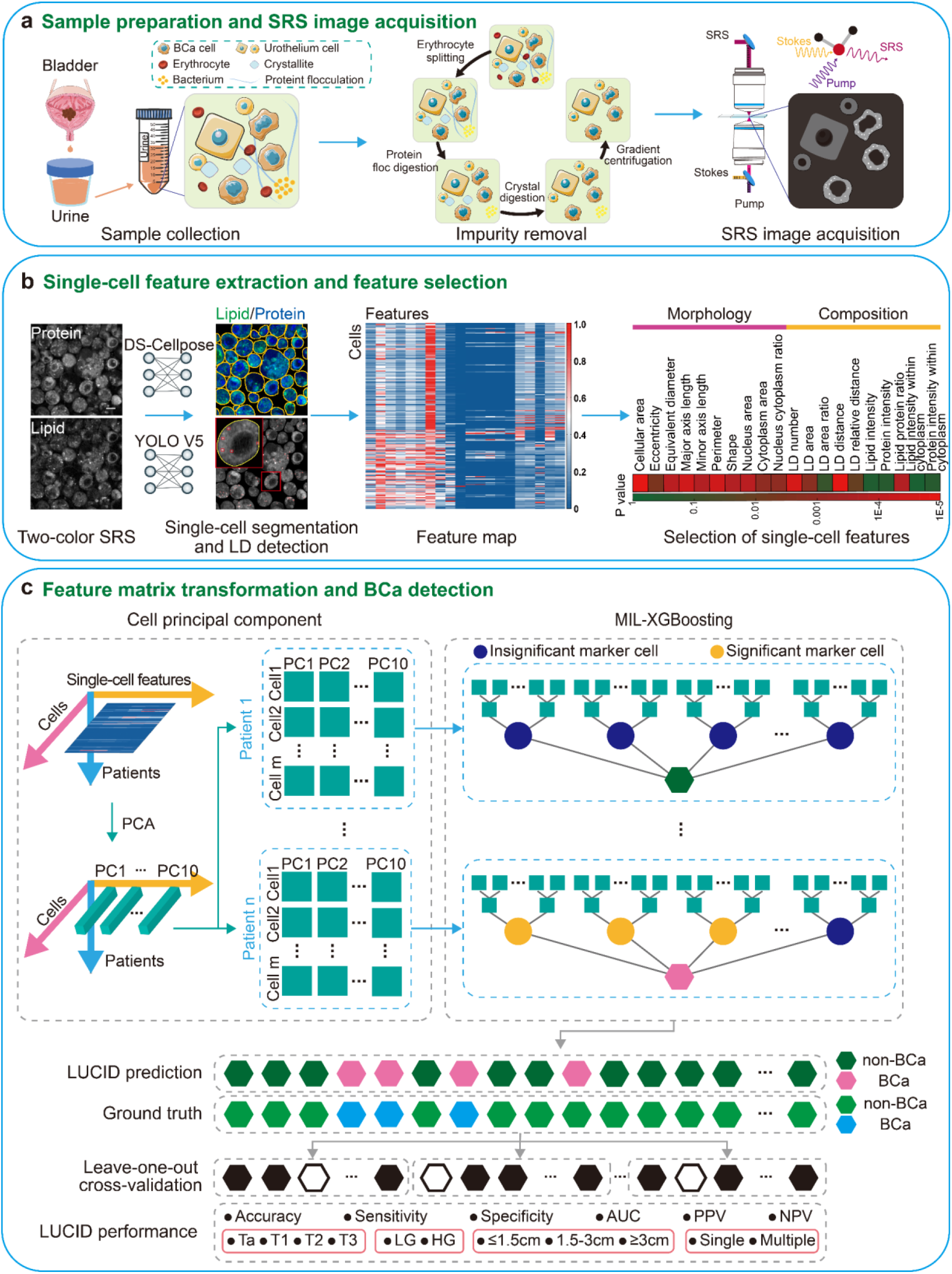
Workflow of the LUCID. **a**, Sample preparation and SRS image acquisition: the workflow consists collection of fresh urine samples, elimination of red blood cells and impurities, and acquisition of SRS images of protein and lipids channels. **b**, Single-cell feature extraction and feature selection: the workflow includes single cell segmentation using the DS-Cellpose model, LD detection using the YOLOv5 model, and extraction and selection of single-cell feature map comprising morphology and composition features. **c**, Feature matrix transformation and BCa detection: the workflow is composed of dimension reduction of feature map into PCs using PCA, recognition of the significant marker cell, serving as the BCa indicator, by using MIL, and BCa diagnosis using the XGBoost model. The performance of LUCID for diagnosis of BCa with different stages, grades, sizes, and numbers, was evaluated by leave-one-out cross-validation, in which the pathological result served as the gold standard.

## Results

### Urine sample preprocessing and SRS imaging

Major impurities in raw urine, including red blood cells, protein flocculates, crystals, and bacteria, can interfere with SRS imaging and subsequent single-cell analysis. To enable reliable SRS imaging of UECs, we developed a stepwise urine preprocessing pipeline to remove these four types of impurities. First, red blood cells not only physically obscured UECs but also generated strong hemoglobin-associated SRS signals within the C–H region (2800–3050 cm^-1^), which can interfere lipid and protein channel quantification. We therefore applied hypotonic lysis followed by washing to remove red blood cells and residual hemoglobin. Second, since collagen -based protein flocculates induced UECs aggregation into clusters that severely impeded single-cell analysis, we employed type I collagenase (37 °C, 1 mg/mL, 10 min) to degrade the collagen network, thereby releasing and dispersing the cells. Third, the urine samples were further treated with 0.1 M NaHCO_3_ to dissolve crystalline components, including calcium oxalate, urate, and phosphate crystals, which can generate strong characteristic Raman signals and introduce substantial spectral interference. Finally, since bacteria (0.5–2 μm) produced Raman signals that overlapped with those of UECs and thereby reduced the signal-to-noise ratio, low-speed centrifugation was employed to reduce free bacteria by taking advantage of their smaller size and lower sedimentation efficiency relative to UECs. Collectively, this preprocessing procedure enriched UECs and reduced major sources of background interference, thereby facilitating subsequent SRS imaging.

After urine sample preprocessing, two-color SRS images of UECs were acquired at the CH_2_-rich band near 2850 cm^-1^ and the CH_3_-rich band near 2930 cm^-1^. Because the SRS spectra of biomolecules exhibit intrinsic spectral overlap, signals acquired at these two Raman bands cannot be attributed exclusively to lipids or proteins. To obtain lipid and protein images, we applied a matrix-unmixing algorithm to resolve the respective molecular contributions. Specifically, because SRS signal intensity is approximately proportional to molecular concentration, the signal measured at each Raman band can be considered a linear superposition of contributions from different molecular components. Following this principle, reference signals obtained from pure chemical standards were used to determine the relative contributions of lipids and proteins and to computationally unmix the two measured SRS channels, as described in detail in the *Methods* section. In summary, following urine preprocessing and two-color SRS imaging, the matrix-unmixing procedure decomposed the original two-color SRS images into lipid (green) and protein (blue) channels for subsequent single-cell analysis.

We next collected urine samples from a primary diagnostic cohort of 80 participants, including 26 patients with BCa and 54 non-BCa participants. The non-BCa cohort consisted of 6 healthy donors and 48 patients with benign urological diseases, including urinary stones (n = 4), interstitial cystitis (n = 3), urinary incontinence (n = 4), and benign prostatic hyperplasia (BPH; n = 37). Detailed clinical characteristics are provided in **Supplementary Table 1**. Following the preprocessing and SRS imaging procedures described above, a total of 867 images were acquired from these participants for subsequent single-cell feature analysis and BCa classification.

### Single-cell Segmentation and Feature Extraction

The detailed steps of single-cell segmentation and feature extraction were shown in **Fig. 2a**. First, we used the protein channel images for single cell segmentation, as the broad intracellular distribution of proteins delineates cell boundaries and morphology more accurately than the lipid channel. To overcome the substantial variation in UEC size and morphology, we developed DS-Cellpose, a modified Cellpose framework that introduces an auxiliary deep supervision branch during the upsampling process to enhance multi-scale segmentation while maintaining a lightweight model architecture. Subsequently, we defined nucleus and cytoplasm masks within each cell by applying an intensity threshold to the lipid channel, as the nucleus contains minimal lipids and consequently exhibits a lower signal intensity, allowing for the quantification of morphological features. In parallel, we utilized the YOLOv5 model to detect LDs from the lipid channel, where LDs are characterized by high signal intensity, appearing as bright small spots. Finally, by integrating these segmentation masks with both the lipid and protein channel images, we comprehensively quantified cellular compositional features.

**Fig. 2.**
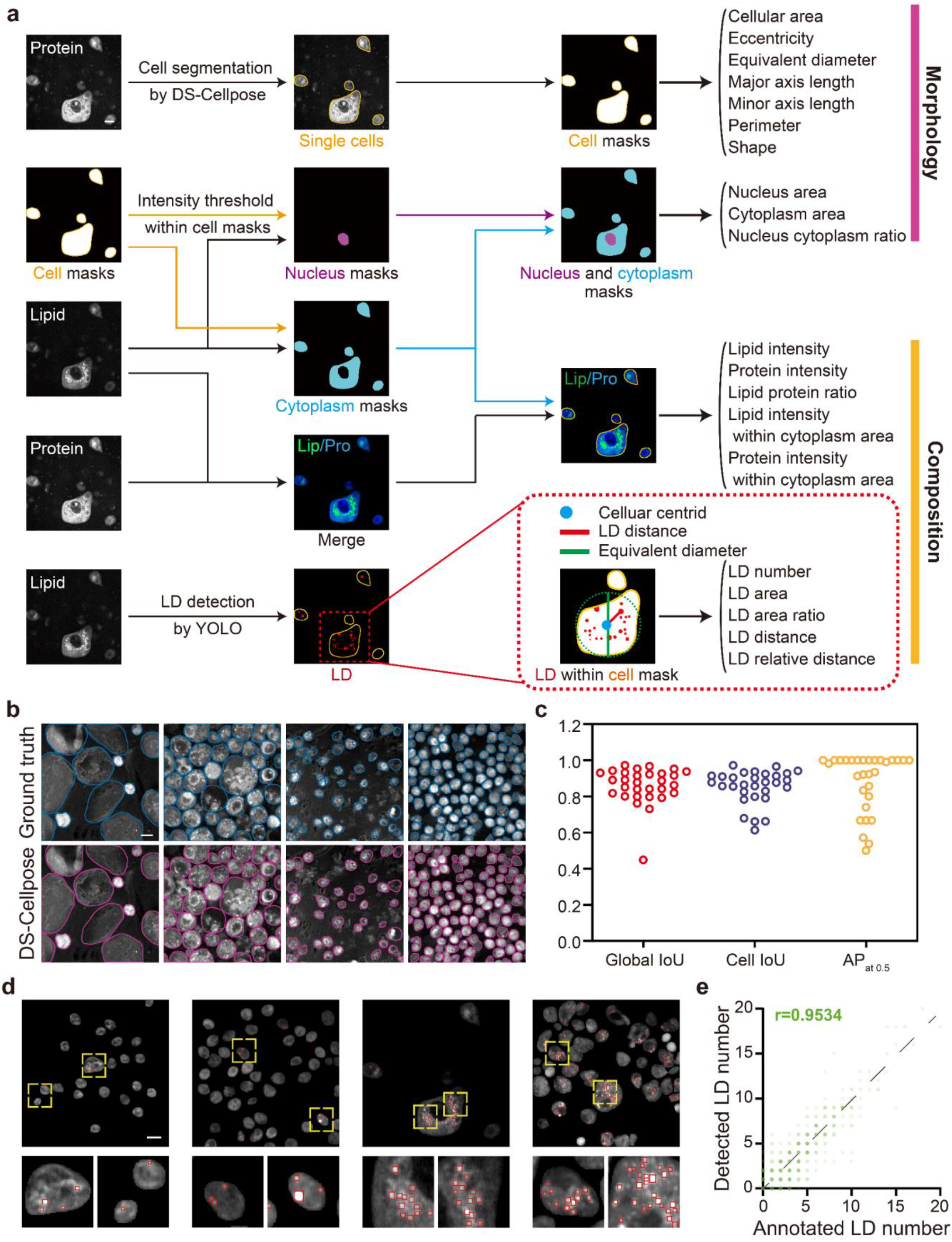
Single-cell segmentation and feature extraction. **a**, Procedures of single-cell segmentation and feature extraction from two-color SRS images. Single-cell features include both morphological and compositional features. **b**, Representative single-cell segmented SRS images by using DS-Cellpose model (purple contour) and the corresponding ground truth (blue contour). **c**, Evaluation of the DS-Cellpose model’s performance in single-cell segmentation, measured by global IoU, cell IoU, and AP_at 0.5_ across 30 images. **d**, Detection of LD (red contour) by the YOLO model. To facilitate visualization, zoom-in images of the representative areas (as indicated by yellow boxes) are shown. **e**, Performance evaluation of the YOLO model for LD detection, based on the consistency of LD number across different cell samples. 3194 cells from five BCa and non-BCa samples were included. *r* represents Pearson correlation coefficient. Scale bars, 10 μm.

For single-cell segmentation, the large variations in the size and shape of UECs make it fundamentally challenging to achieve robust performance using conventional algorithms, such as Cellpose, StarDist, U-Net, and Watershed. As shown in **Supplementary Fig. 1a**, Watershed frequently produced over-segmentation in regions with heterogeneous intracellular intensity, whereas U-Net and StarDist showed limitations in separating adjacent or morphologically irregular cells. Although Cellpose worked better than the other three methods, its segmentation accuracy decreased when segmenting cells with significant size variations within the same image. To address these challenges, we developed DS-Cellpose by integrating a Deep Supervision (DS) module into the Cellpose framework, thereby improving segmentation performance (**Supplementary Fig. 1a**). As shown in **Fig. 2b&c**, DS-Cellpose achieved a Global IoU of 0.86 (reflecting overall foreground coverage), a Cell IoU of 0.85 (indicating the precision of individual instance separation), and an average precision (AP_at 0.5_) of 0.88. As shown in **Supplementary Fig. 1c**, compared to Cellpose, DS-Cellpose showed higher Global IoU (0.86 vs. 0.78), Cell IoU (0.85 vs. 0.74, p < 0.01), and AP_at 0.5_ (0.88 vs. 0.68, p < 0.01), and significantly outperformed all other methods across these metrics.

To quantitatively evaluate the performance of DS-Cellpose in scenarios exhibiting strong morphological heterogeneity, we respectively analyzed segmentation performance for big (diameter ≥ 12.5 µm) vs. small (diameter < 12.5 µm), as well as regular (shape ≥ 0.88) vs. irregular (shape < 0.88) cells, which were categorized based on the distinct distributions (**Supplementary Fig. 1b**). As shown in Supplementary Fig. 1c, DS-Cellpose consistently outperformed Cellpose across all morphological subtypes and showed significant improvements in Global IoU, Cell IoU, and AP_at 0.5_ compared with the other three methods. Notably, when compared specifically to Cellpose, DS-Cellpose exhibited superior robustness in handling big or irregular cells, achieving significantly higher metrics for big & irregular cells (Cell IoU: 0.81 vs. 0.59; AP_at 0.5_: 0.77 vs. 0.50), big & regular cells (AP_at 0.5_: 0.98 vs. 0.81), and small & irregular cells (AP_at 0.5_: 0.91 vs. 0.72). This indicates that DS-Cellpose, through its deep supervision branch, enhanced its multi-scale segmentation capability, thereby better addressing the challenge posed by morphological variations among UECs within a single image. Collectively, these results indicate that DS-Cellpose provides robust single-cell segmentation across UECs with substantial variation in size and morphology.

Then, by integrating the single cell mask with the two-color SRS image, totally 20 single-cell features of UECs were extracted, including 10 morphological features (cellular area, eccentricity, equivalent diameter, major axis length, minor axis length, perimeter, shape, nucleus area, cytoplasm area, and nucleus-cytoplasm ratio) and 10 compositional features (lipid intensity, protein intensity, lipid-protein ratio, lipid intensity within cytoplasm area, protein intensity within cytoplasm area, and 5 LD-related features). By integrating the LD masks in single cells, 5 LD-related single-cell features were extracted, including LD number, LD area, LD area ratio, LD distance, and LD relative distance.

To enable robust quantification of LD-associated features, we developed a YOLOv5-based detector using lipid-channel SRS images (**Fig. 2d** and **Supplementary Fig. 2a**). The Laplacian of Gaussian (LoG) operator, a traditional blob detection method, served as the baseline. Specifically, lipid channel SRS images were fed into the YOLO model to extract LDs according to their dotted shapes and intensity maxima. Its detection results conformed well to the LDs shown in the lipid channel SRS images. In contrast, LoG required frequent manual setting of parameters such as maxima threshold and kernel size to adapt to data variability, impeding accurate, automated, and objective LDs detection. The correlation coefficient between the number of LDs detected by the YOLO model and in the manual annotation was 0.95, which was much higher than 0.15 for LoG with fixed parameters (**Fig. 2e, Supplementary Fig. 2b**). The difference in LD number between BCa and non-BCa samples detected by YOLOv5 showed an effect size comparable to that obtained using manual annotations (η^2^ = 0.1635 vs. 0.1493), whereas the LoG-derived difference showed a substantially smaller effect size (η^2^ = 0.0139; **Supplementary Fig. 2c**). Based on the YOLO model, 5 LD-related single-cell features were extracted, including LD number, LD area, LD area ratio (the ratio of LD area to cellular area), LD distance (the mean distance between LDs and cellular centroid), and LD relative distance (the ratio of LD distance to cellular equivalent diameter). The detailed procedure of feature extraction is described in the *Methods* section.

### Feature Comparisons within Cell Lines

In order to identify distinct features of BCa samples, we first analyzed SRS images of cultured cell lines, including normal urothelium SV-HUC-1 and three BCa lines (BIU-87, 5637, and T24) (**Supplementary Fig. 3**). Six features with significant differences and low inter-feature correlation were selected: three morphological features (cellular area, nucleus area, nucleus-cytoplasm ratio) and three compositional features (LD area ratio, LD distance, lipid intensity). Logistic regression classification using these six features was able to distinguish BCa cells from normal cells. Specifically, distinguishing each cancer cell line from SV-HUC-1 yielded optimal AUCs of 0.93 for BIU-87 (using compositional features), 0.99 for 5637 (using combined morphological and compositional features), and 0.93 for T24 (using morphological features). Collectively, these results suggest that despite the variation in dominant features across BCa cell lines, BCa cells may have distinctive morphological and/or compositional features, which could be used for differentiation from non-BCa.

### Differential Single-cell Features Between BCa and Non-BCa

We extracted single-cell features from totally 19,678 UECs in non-BCa cohort (n = 54) and BCa cohort (n = 26). The features were quantified for each donor or patient and shown in a heatmap (**Fig. 3a**). Totally 15 features were found to be significantly different between non-BCa and BCa samples (**Fig. 3b, Supplementary Table 2**). Specifically, the cell size and nucleus relevant features, including cellular area, equivalent diameter, major axis length, minor axis length, perimeter, cytoplasm area, nucleus area, and nucleus cytoplasm ratio, were significantly larger in BCa samples compared to non-BCa ones. In contrast, cell circularity relevant features, such as shape, showed a marked decrease in BCa samples. As shown in **Supplementary Fig. 4**, similar results were obtained from statistical analysis of single-cell features in non-BCa and BCa cases. Notably, a majority of LD features, including LD number, LD distance, LD area, and LD area ratio, were remarkably elevated in BCa samples. This finding revealed aberrant LD accumulation in BCa UECs, which is consistent with previous studies on dysregulated lipid metabolism in BCa^20,22,23^.

**Fig. 3.**
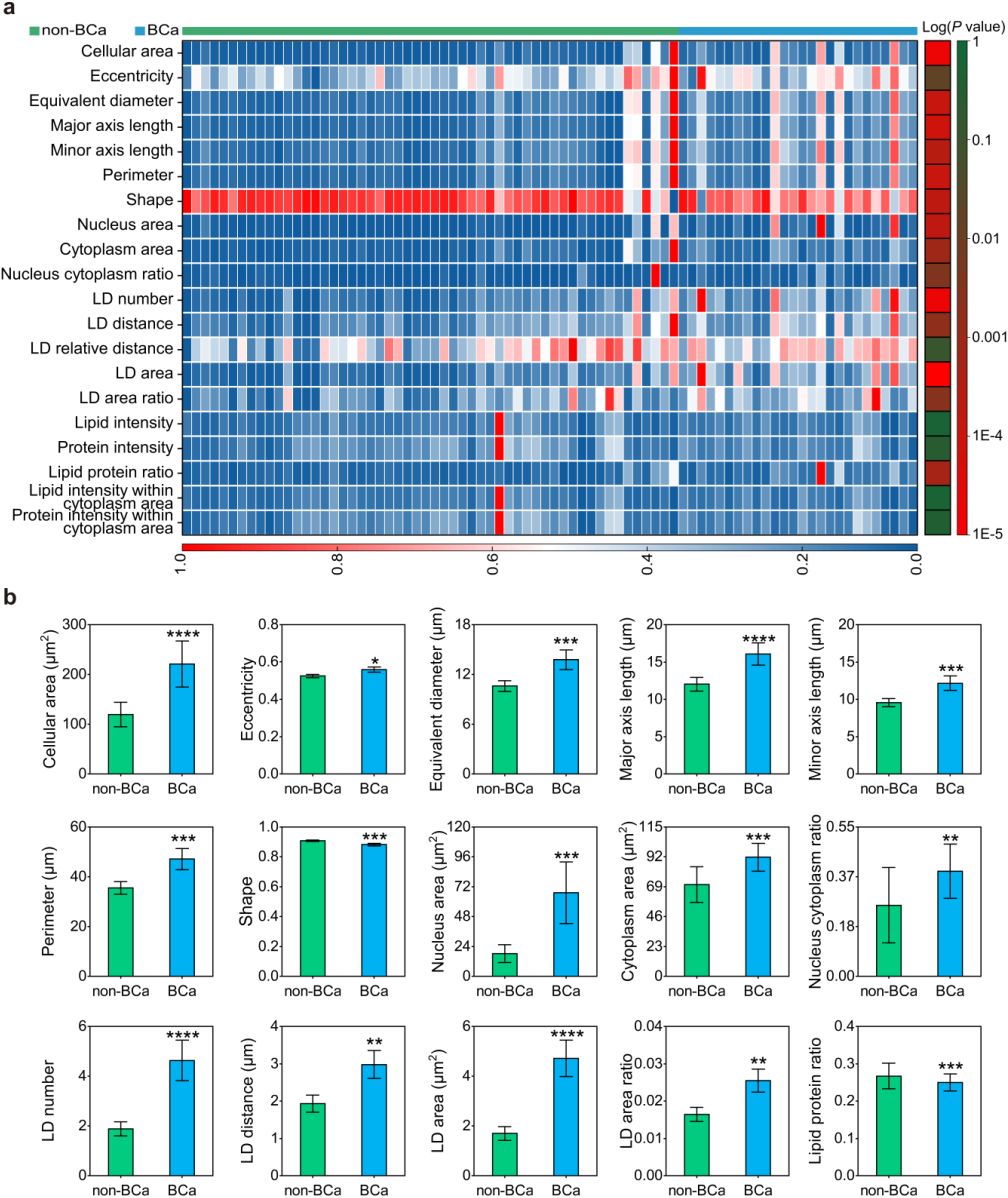
Differential single-cell features between non-BCa and BCa. **a**, Heatmap and comparison of 20 single-cell features between non-BCa (n = 54 donors/patients) and BCa (n = 26 patients), where the column of *p* values was obtained by Mann-Whitney U test. **b**, Comparison of the 15 features that differed significantly between the two groups. Bars represent group means, and error bars indicate the standard deviation (SD). Statistical significance is indicated by the *p* value from the Mann-Whitney U test (ns: non-significant, *: *p* <0.05, **: *p* <0.01, ***: *p* <0.001, ****: *p* <0.0001).

### LUCID Enables Precise BCa Diagnosis via Lipid Droplet Features

Given the similarity in the types of UECs between BCa and non-BCa, primarily consisting of benign epithelial cells, it is essential to identify significant marker cells with specific features that can precisely indicate BCa. To address this, we developed a MIL-XGBoosting algorithm, which treats each patient as a bag composed of individual cellular instances represented by 10 PCs derived from PCA dimensionality reduction, and employs XGBoost as a classifier to identify significant marker cells serving as indicators of BCa. Notably, a cell is classified as a significant marker cell when its predicted positive probability exceeds 0.01. Consequently, the positive probability for each patient is defined as the maximum predicted probability observed among the constituent cells.

To validate the diagnostic performance of the MIL-XGBoosting algorithm within the LUCID framework, we established a cohort of 80 patients (26 BCa and 54 non-BCa), comprising a total of 19,678 UECs. Employing leave-one-out cross-validation, we conducted feature-ablation experiments to assess the contributions of different cellular feature sets to BCa classification. This involved comparing individual feature sets (LD, morphology, and Lipid & Protein), as well as their comprehensive integrations (e.g., morphology & LD; **Fig. 4a, b**). Owing to the intrinsic morphological heterogeneity of UECs, morphology alone showed limited discriminative performance (AUC: 0.739; accuracy: 70.00%; specificity: 75.93%; sensitivity: 57.69%), resulting in substantial false positives (PPV: 53.57%). In contrast, compositional features, particularly LD features, provided stronger discriminative information, with AUC values of 0.863 for complete compositional features and 0.887 for LD features alone. Notably, compared with Lipid & Protein features alone, adding LD features to form the complete compositional feature set increased sensitivity from 46.15% to 65.38%, specificity from 77.78% to 90.74%, accuracy from 67.50% to 82.50%, PPV from 50.00% to 77.27%, and AUC from 0.593 to 0.863 (**Fig. 4b**), highlighting the substantial contribution of LD features to BCa discrimination. Furthermore, the integration of different feature sets revealed complementary contributions to BCa identification. Adding morphology to LD increased sensitivity from 80.77% to 84.62% while maintaining the same specificity of 77.78%, whereas incorporation of Lipid & Protein into LD increased specificity from 77.78% to 90.74%, accompanied by a decrease in sensitivity from 80.77% to 65.38%. Importantly, integrating morphology with composition (LD + Lipid & Protein) further increased sensitivity from 65.38% to 92.31% and AUC from 0.863 to 0.940, while maintaining a high specificity of 87.04% (**Fig. 4b**). Overall, feature-ablation analysis identified LD-associated features as a major source of discriminative information, while their integration with broader compositional and morphological features jointly contributed to high diagnostic sensitivity and specificity.

**Fig. 4.**
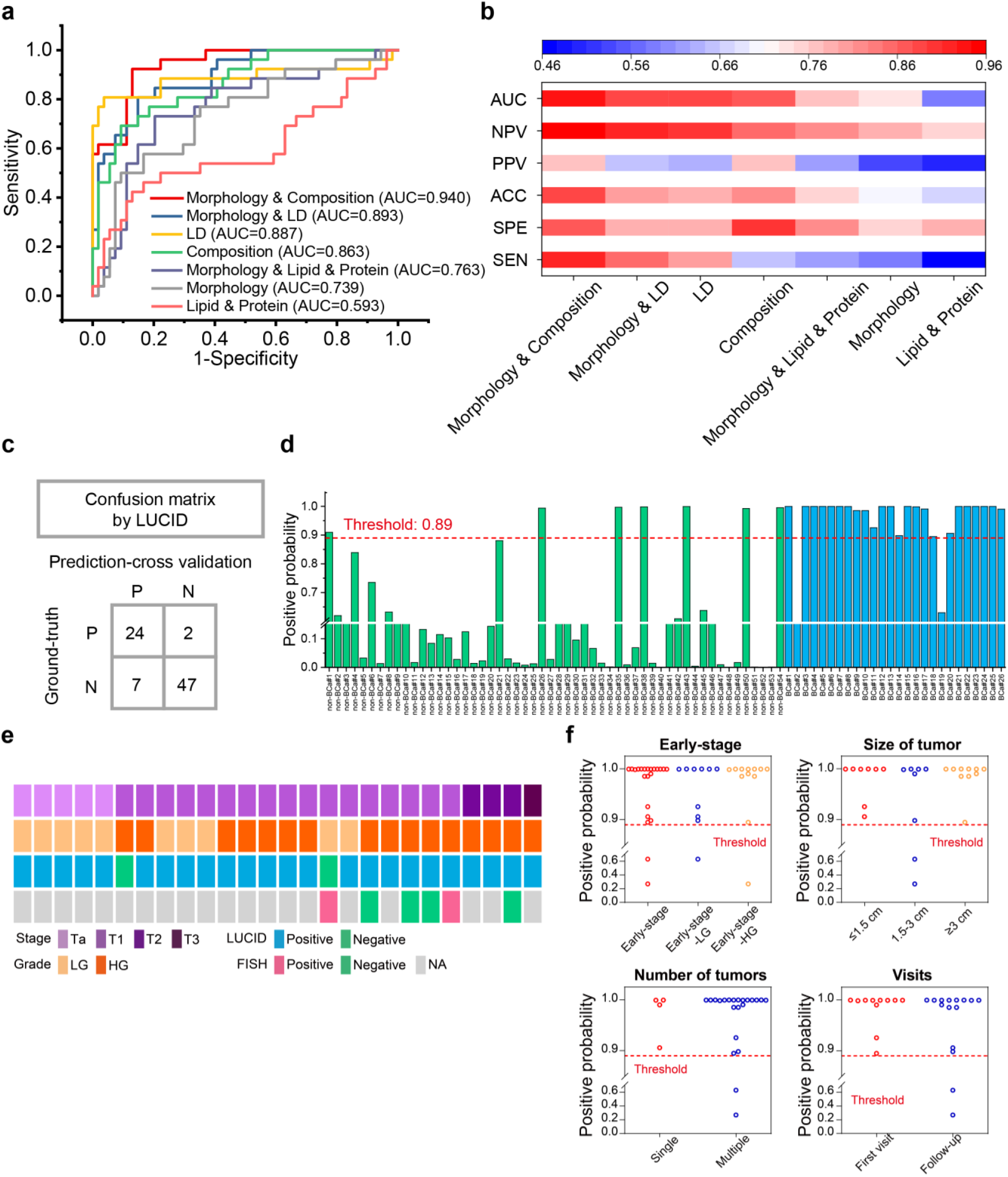
Lipid droplet features drive the precise BCa diagnosis in the LUCID framework. **a**, Comparison of receiver operating characteristic (ROC) curves and AUC of BCa detection with different feature combinations (LD features, morphology features, morphology features & LD features, composition features, morphology & composition features, lipid & protein features, morphology & lipid & protein features). **b**, Comparison of performance metric values of BCa detection with different feature combinations. **c**, Confusion matrix of cross validation in BCa detection by LUCID. **d**, Positive probability of BCa detection by LUCID. Red dashed line, threshold (0.89) of positive prediction. **e**, Clinical features of BCa patients (staging and grading) and their corresponding LUCID prediction results, along with FISH testing results. **f**, Quantitative accuracy of LUCID predictions based on different tumor clinical feature classifications (including early-stage, tumor size, tumor number, and number of visits). Each hollow circle represents the positive prediction value for a single patient. The red dashed line indicates the positive prediction threshold (0.89).

To further interpret these results, we performed a single-cell-level analysis of the feature space in the patient cohort dataset. First, feature importance analysis showed that PC1 contributed more strongly than the other PCs to the classifier (**Supplementary Fig. 13a**). As shown in PC1–PC2 plot, the non-BCa cells exhibited a highly concentrated distribution, forming a compact, high-density core, whereas the BCa population displayed a markedly broader and more diffuse distribution that overlapped substantially with the non-BCa population (**Supplementary Fig. 12**). The significant marker cells were preferentially located in the more dispersed regions of the BCa population and tended to exhibit higher PC1 and PC2 values, indicating that the model identified a specific subpopulation of BCa-associated cells rather than the entire BCa population. Furthermore, as shown in **Supplementary Fig. 15**, although significant marker cells were also present in non-BCa samples, their marker-cell probabilities were significantly lower than those of significant marker cells in BCa samples.

As illustrated in the patient-level probability bar chart (**Fig. 4d**), this identification of significant marker cells translated into high diagnostic confidence, with correctly predicted BCa patients generally exhibiting high positive probabilities, whereas most non-BCa participants showed substantially lower positive probabilities. Moreover, comparative benchmarking against other MIL classifiers revealed that XGBoosting exhibited higher diagnostic performance than Support Vector Machine (SVM) and Logistic Regression (LR) in the evaluated MIL framework (**Supplementary Fig. 11, Supplementary Table. 3**). Collectively, these results demonstrate that LUCID captures the heterogeneous cellular characteristics of BCa and identifies significant marker cells enriched in the higher-PC1/PC2 feature space, thereby enabling precise and robust BCa diagnosis.

To further elucidate the cellular characteristics underlying the diagnostic performance of the MIL-XGBoosting algorithm, we performed correlation analyses to interpret the contribution of individual cellular features. Correlation analysis showed that PC1 was strongly associated with multiple morphological and LD-associated features, suggesting that variation along PC1 reflected the joint contributions of these cellular characteristics (**Supplementary Fig. 13b**). Notably, although morphological features contributed most strongly to this axis, morphology alone was insufficient for accurate discrimination, as some non-BCa cells exhibited morphological patterns overlapping with those of BCa-associated cells. In contrast, integration with compositional features, particularly LD-related features, provided additional discriminative information for distinguishing these overlapping cellular populations. As shown in **Supplementary Fig. 14a**, both morphological features (e.g., Cellular Area, Shape) and LD-related features (e.g., LD number, LD Area) exhibited significant correlations with marker-cell probability (Cellular Area: Pearson r = 0.4366, p < 0.0001; Shape: Pearson r = −0.2353, p < 0.0001; LD number: Pearson r = 0.4863, p < 0.0001; LD Area: Pearson r = 0.5232, p < 0.0001). These results indicate that the model tends to assign higher marker-cell probabilities to cells characterized by larger cellular area, more irregular morphology, and increased LD content. Collectively, these results further support LD features as a major source of discriminative information, while morphological and other compositional features provide complementary information for the identification of BCa-associated marker cells and patient-level classification.

To validate the capability of LUCID for early diagnosis and different tumor clinical feature classifications, we further assessed the concordance between model predictions and pathological diagnoses across diverse dimensions. As shown in **Fig. 4e**, the 26 BCa patients were stratified by clinical stage into Ta (n = 5), T1 (n = 17), T2 (n = 3), and T3 (n = 1), and by tumor grade into low-grade (LG, n = 10) and high-grade (HG, n = 16) cases. Among them, six patients also underwent FISH testing. LUCID achieved 90.91% (20/22) concordance with pathological diagnoses in early-stage tumors (Ta and T1), including 9 of 10 low-grade early-stage tumors (90.0%; **Fig. 4f**). Among the six patients who underwent both assays, LUCID exhibited higher concordance with pathology than FISH (83.33% [5/6] vs. 33.33% [2/6]) (**Fig. 4e**). We further evaluated LUCID’s diagnostic performance across cases with varying tumor sizes and numbers. Notably, LUCID successfully identified all patients presenting with small tumors (≤1.5 cm; 8/8) and single tumors (4/4), demonstrating its high sensitivity for early-stage and minimal BCa lesions (**Fig. 4f**). Moreover, this robust diagnostic accuracy was consistently maintained across different clinical scenarios, achieving 100% (11/11) in the first-visit cohort and 86.67% (13/15) in the follow-up cohort (**Fig. 4f**). Furthermore, we evaluated LUCID on a cohort of 54 non-BCa participants, and it maintained reliable diagnostic performance, with concordance rates of 5/6 for normal bladders, 3/4 for urinary stones, 2/3 for interstitial cystitis, 3/4 for urinary incontinence, and 34/37 for BPH. Collectively, these results highlight LUCID maintained high detection rates across several clinically relevant BCa subgroups while retaining substantial specificity in non-BCa participants.

### Identification of The Significant Marker Cell for BCa

The robust diagnostic performance of LUCID, particularly its high sensitivity in detecting early-stage and small tumors, underscores the efficacy of the MIL-XGBoosting framework in finding specific pathological signatures from a complex background of benign epithelium. Central to this capability is the identification of significant marker cells, as specific cellular instances with high predictive probabilities that drive the patient-level diagnosis. To elucidate the biological basis of these predictions and explain that the model captures biologically meaningful phenotypic changes, we proceeded to characterize these pivotal cells.

To visualize the difference of significant and insignificant marker cells identified by LUCID, we used SRS images from six representative BCa and non-BCa samples (BCa#7 and non-BCa#16, BCa#22 and non-BCa#25, BCa#21 and non-BCa#42) as examples (**Fig. 5a**). Significant marker cells in the BCa samples exhibited larger cellular area, more irregular morphology, and a distinct accumulation of LDs, together with higher predicted probabilities. In contrast, corresponding cells in non-BCa samples, were characterized by smaller cell size, more regular morphology, and lower positive probabilities, although LD accumulation was also observed in some cells. To illustrate the spatial relationship between significant marker cells and insignificant marker cells in the PCA space, we extracted PC1 and PC2 and plotted the corresponding two-dimensional PCA space (**Fig. 5b**). To enhance the clarity of the point distributions, significant marker cells with extreme PC values are excluded from the plot, and the complete data is shown in **Supplementary Fig. 5**. In the PCA scatter plot, significant marker cells (yellow) were predominantly distributed in the more dispersed regions of the BCa population and were characterized by relatively higher PC1 and PC2 values, whereas insignificant cells from both BCa (blue) and non-BCa (green) samples were mainly concentrated within the lower-value region.

**Fig. 5.**
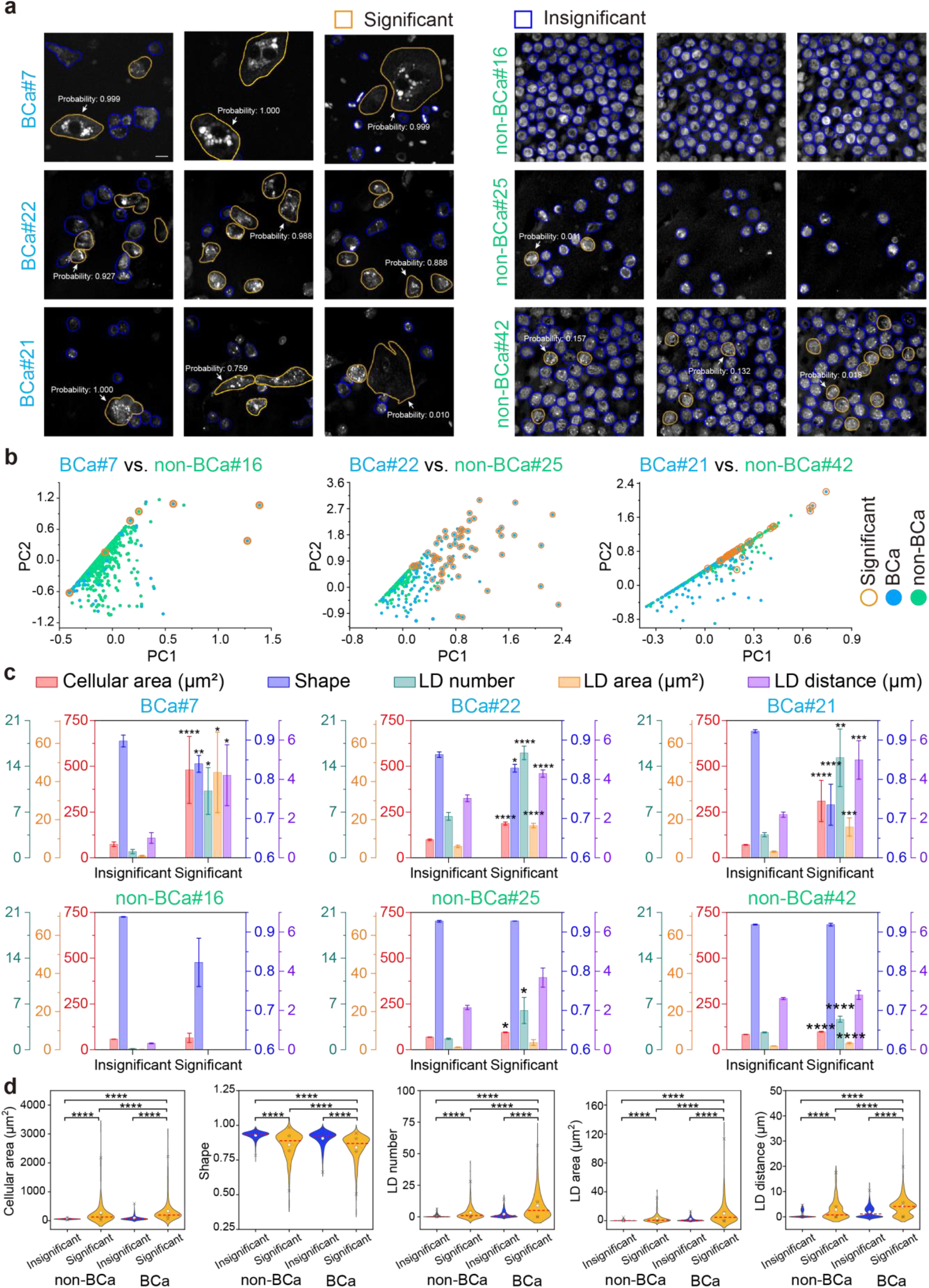
Marker cells identification by LUCID. **a**, Representative SRS images of BCa and non-BCa pairs: BCa#7 and non-BCa#16, BCa#22 and non-BCa#25, BCa#21 and non-BCa#42. Significant marker cells highlighted by yellow contours and insignificant cells by blue contours. Scale bar, 10 μm. **b**, PC1 vs. PC2 plots for sample pairs. Highlighted points denote significant marker cells. Complete scatter plots are provided in Supplementary Fig. 5. **c**, Quantitative comparison of representative cellular features in 6 samples between significant and insignificant marker cells. Bars represent group means, and error bars indicate the standard deviation (SD). **d**, Violin plots of representative cellular features in all non-BCa and BCa samples between significant and insignificant marker cells. Within each violin plot, the symbols and lines denote the following statistical parameters: ×: 1st and 99th percentiles; ☆: 25th and 75th percentiles; ○: mean value; Red dashed lines: median. Statistical significance is indicated by the *P* value from the Mann-Whitney U test (ns: *P* >0.05, *: *P* <0.05, **: *P* <0.01, ***: *P* <0.001, ****: *P* <0.0001).

Quantitative analysis revealed that significant marker cells exhibited distinct compositional and morphological characteristics compared with insignificant cells across the representative BCa samples, with significantly increased cellular area and LD content, accompanied by a decrease in shape factors (**Fig. 5c**). These differences were observed across multiple parameters, reflecting distinct intracellular accumulation patterns consistent with the SRS images (**Fig. 5a**). Similar trends were observed across the corresponding samples, with significant marker cells showing consistently higher LD feature values and cellular area, but lower shape factor values than insignificant cells (**Fig. 5d**). Consistently, as shown in Cellular area-LD area plot (**Supplementary Fig. 14b**), significant marker cells were enriched in regions characterized by increased LD content and larger cell size, whereas non-significant cells were mainly confined to a compact low-value region. Quantitative analysis of other features is presented in **Supplementary Fig. 6** and **Supplementary Fig. 7**. Collectively, these results highlight the importance of LD-related and morphological features in characterizing significant marker cells for BCa. Significant marker cells exhibiting prominent LD accumulation, larger cellular area, and more irregular morphology could be identified with high confidence in BCa samples, further supporting the potential of these cellular features as diagnostic markers for precise and non-invasive BCa detection.

Next, we examined the correlation between these significant marker cells identified by MIL-XGBoosting and the tumor cells annotated by cytologists on Hematoxylin and Eosin (H&E) staining. Using an additional cohort of four cases (BCa #27, BCa #28, non-BCa #55, and non-BCa #56), we compared differences in single-cell SRS and *in situ* H&E cytological images features between BCa and non-BCa samples (**Supplementary Fig. 16**). Although the significant marker cells identified by MIL-XGBoosting on SRS images did not fully coincide with the tumor cells annotated by cytologists on in situ H&E images, the significant marker cell population encompassed the annotated tumor cells in BCa samples and exhibited strong consistency in non-BCa samples, where neither significant marker cells nor annotated tumor cells were detected (**Supplementary Fig. 16a**). Notably, LD features differed significantly between the tumor and normal cells annotated by cytologists, consistent with the differences observed between significant and insignificant marker cells identified by MIL-XGBoosting (**Supplementary Fig. 16b**). In particular, both LD area and LD number showed significant increases, consistent with the preceding analyses. These findings suggest that BCa detection based on LD feature alterations is consistent with conventional cytological morphology, while the broader distribution of these significant marker cells reveals that metabolic alterations may capture diagnostically relevant changes beyond those identified by H&E-based morphological criteria alone. Collectively, the LUCID framework integrates morphological and compositional information, including LD features, and thereby provides enhanced sensitivity for BCa detection compared with conventional morphology-based cytology.

## Discussion

In this study, we developed LUCID, a non-invasive molecular cytology framework that integrates label-free SRS microscopy with machine learning for BCa detection using urinary exfoliated cells. Compared to conventional cytology, which involves complex staining procedures and relies on morphological criteria, our LUCID method enables non - invasive, rapid imaging of UECs while simultaneously acquiring both compositional and morphological information. By further leveraging LD-related compositional features, it significantly enhances the diagnostic sensitivity for BCa. The following discussion demonstrates that LUCID may open up a new avenue for the accurate detection of BCa.

First, from a clinical perspective, LUCID shows strong potential for improving the non-invasive detection of BCa by combining high diagnostic sensitivity with label-free and non-destructive cellular analysis. Effective early BCa diagnosis and surveillance are critical for reducing BCa mortality, owing to the disease’s high incidence and recurrence rates, which necessitate frequent, rigorous follow-ups^36^. While non-invasive liquid biopsies based on UECs present a promising alternative to cystoscopy, current methods consistently fail to achieve adequate diagnostic sensitivity, particularly for early-stage and small tumors^37,38^. Compared with conventional urine cytology (sensitivity ∼40%)^6,7^, FISH (sensitivity 65%, specificity 81%)^9,39,40^, and methylation testing (sensitivity 90%, specificity 83%)^11^, our LUCID increases sensitivity to 92.31%, specificity to 87.04%, and overall accuracy to 88.75% in our cohort, and notably showed 90.91% concordance with pathological diagnoses in early-stage BCa (**Supplementary Table 4**). Moreover, its label-free and non-destructive nature may preserve cellular specimens for subsequent molecular or genomic analyses. Notably, among the patients with BCa in this study, six underwent both LUCID and FISH testing; LUCID was positive in 5 of 6 cases, whereas FISH was positive in 2 of 6 cases (**Fig. 4e**). Further histopathological support was provided by in situ H&E validation, in which pathologist-annotated tumor cells in BCa samples corresponded to LUCID-identified significant marker cells, whereas non-BCa samples consistently showed neither significant marker cells nor tumor cells (**Supplementary Fig. 16**). Together, these findings indicate that LUCID can sensitively identify BCa-associated cells while preserving specimens for downstream analyses, supporting its potential as a complementary approach for non-invasive BCa detection.

Second, LUCID establishes a robust technical pipeline for SRS-based urine cytological analysis by addressing several key challenges inherent to urinary exfoliated cells. Complex urinary components, including red blood cells, urinary crystals, proteinaceous flocculent material, and bacteria, which may compromise the diagnostic accuracy in traditional cytology^41,42^. To address this, we established a standardized urine sample preparation protocol to reduce these impurities and enrich UECs for subsequent SRS imaging. In addition, the pronounced morphological heterogeneity of UECs, including BCa cells, squamous epithelial cells, urothelial cells, and immune cells^43-45^, complicates single-cell segmentation and feature extraction. For single-cell segmentation, we developed DS-Cellpose, an improved Cellpose^46^ framework that introduces a deep supervision branch within the decoder to enhance multi-scale segmentation capability, outperforming other methods across cells with diverse morphologies (**Supplementary Fig. 1**). We further implemented a YOLO-based method using the YOLOv5^47^ model to identify LDs directly from the lipid channel, enabling more accurate extraction of LD-related information than the LoG-based method. This approach not only ensures accurate extraction of lipid droplet information from UECs, but also provides a solution to the challenge of LD detection in SRS image processing. To further address the low proportion of malignant UECs and the absence of cell-level labels, we developed a hybrid PCA and MIL-XGBoosting framework that identifies rare BCa-associated marker cells and achieved 92.31% sensitivity, 87.04% specificity, and an AUC of 0.940 (n = 80). Compared with average pooling, the MIL strategy better preserved diagnostically informative rare-cell signals (**Supplementary Fig. 17**), while feature-ablation analysis further showed that LD-associated features provided substantial discriminative information and that their integration with broader compositional and morphological features yielded the best overall performance (**Fig. 4a, b**). Collectively, these technical advances address challenges in sample preparation, heterogeneous cell segmentation, LD detection, and rare-cell classification, providing a robust technical pathway for SRS-based urine cytological analysis.

Third, examining the significant marker cells identified by LUCID revealed that LD features serve as the core discriminative markers for BCa. As shown in **Supplementary Fig. 6** and **Supplementary Fig. 7**, the significant marker cells identified by LUCID exhibited significantly elevated LD feature values at both the single-cell and patient levels. From a molecular biology perspective, BCa tissues commonly exhibit upregulation of the PI3K– AKT–mTOR pathway and inhibition of AMPK, which enhances acetyl-CoA carboxylase activity, activates fatty acid synthesis, and leads to the storage of neutral lipids in the form of LDs within bladder cancer cells^19,20,48^. LDs are closely associated with BCa survival^49^, invasion^50^, and metastasis^51^. Moreover, recent studies have also identified lipid-related urinary biomarkers for BCa. For example, urinary neolactotetraosylceramide (nLc4), a tumor-associated glycosphingolipid, has been reported as a potential biomarker for MIBC detection^50^. Furthermore, as shown in **Supplementary Fig. 8**, the LD feature values of significant marker cells were higher than those of other cells in the corresponding cases across all 80 patients, and a substantial proportion of these marker cells were detected in all correctly classified BCa samples (**Supplementary Fig. 9**), indicating that elevated LD features serve as a reliable marker for BCa rather than an artifact driven by a few patients.

Finally, from a translational perspective, LUCID shows great promise for improving BCa liquid biopsy diagnostics, but several challenges remain to be resolved. Among the nine misclassified samples (**Supplementary Fig. 10**), one high-grade BCa case contained only six cells (BCa#02), which may have increased the likelihood of missing diagnostically informative cells, whereas the other misclassified BCa case was low-grade and retained a relatively high patient-level probability compared with most non-BCa samples. The seven misclassified non-BCa samples contained only a small number of significant marker cells. These observations suggest that automated whole-slide scanning and hyperspectral or super-resolution SRS microscopy may improve cell capture and single-cell characterization, thereby reducing misclassification. In addition, the current model was evaluated in an 80-patient cohort using leave-one-out cross-validation, which provides a reliable estimate of internal generalization but does not replace rigorous external validation; therefore, large-scale, independent, multicenter prospective cohorts are still required to establish the robustness and generalizability of LUCID. Direct comparisons with standard-of-care methods also remain limited, as concurrent LUCID and FISH testing was performed in only six patients and in situ H&E validation in four additional cases. Larger head-to-head studies are therefore needed to more rigorously define the clinical performance of LUCID. Further development of an end-to-end architecture may also improve robustness and clinical deploy ability by reducing error propagation across sequential processing steps

## Methods

### Cell culture

BIU-87 and 5637 cells were cultured in RPMI 1640 medium (Gibco, 11875119), T24 cells were cultured in McCoy’s 5A medium (Gibco, 16600082), and SV-HUC-1 cells were cultured in Ham’s F-12K medium (Gibco, 21127022). All media were supplemented with 10% (v/v) fetal bovine serum (Omega Scientific, BF-21) and 0.1% (v/v) penicillin-streptomycin (Gibco, 15070063) throughout culture. Cell cultures were maintained in a humidified incubator at 37 °C in 5% CO^2^.

### Clinical sample

Preoperative morning urine samples were collected from 80 participants at Peking University First Hospital, including 26 BCa and 54 non-BCa patients. Clinical information, including age, sex, control type, tumor stage, tumor grade, tumor size, and single/multiple tumor occurrence, was provided in **Supplementary Table 1**. This study was approved by the Institutional Review Boards of Beijing Cancer Hospital and Beihang University, and written informed consent was obtained from all participants.

### Preparation of UEC for SRS imaging

For each patient, 40 mL of midstream morning urine was collected and centrifuged at 2000 rpm for 5 min. The supernatant was discarded, leaving ∼0.5 mL. Red blood cell lysis buffer was added, and the mixture was incubated in an ice–water bath for 10 min, followed by brief vortex mixing. After centrifugation (2000 rpm, 5 min), the supernatant was removed, leaving ∼0.1 mL. The pellet was treated with 0.5 mL of a solution containing type I collagenase (1 mg/mL) and 0.1 M NaHCO_3_ and incubated at 37 °C for 10 min. Following another centrifugation step (2000 rpm, 5 min), the supernatant was discarded, leaving ∼0.1 mL. Cells were resuspended in PBS to a final volume of 1 mL and centrifuged at 500 rpm for 5 min. The supernatant was removed, leaving ∼0.1 mL, and the cell concentration was adjusted to 1 × 10^7^ cells/mL. A 3 µL aliquot of the suspension was spread evenly (∼0.3 cm^2^) on a glass slide, and air-dried for SRS imaging. Samples were stored at −80 °C until use.

### Two-color SRS microscopy

SRS microscope employed a dual-output picosecond pulse laser (picoEmerald™ S, Applied Physics and Electronics) with a repetition rate at 80 MHz and 2 ps pulse width. The laser outputted spatiotemporally overlapped pump beam with tunable wavelength from 700 nm to 960 nm and the Stokes beam with fixed wavelength at 1031 nm. When performing SRS, the Stokes beam was modulated at ∼20 MHz by an electronic optic modulator. The collinear pump and Stokes beams were coupled to a 2D scanning galvanometer (GVS012-2D, Thorlabs) and then imported into an inverted microscope (IX73, Olympus). A 60× water immersion objective lens (LUMPlanFL N, 1.0 numerical aperture, Olympus) focused the lasers into the sample. The photons were then collected by a 60× water-immersion objective (LUMPlanFL N, 1.0 numerical aperture, Olympus). The pump beam was selected by a short-pass filter (ET980SP, Chroma) and detected by a photodiode (S3994-01, Hamamatsu, Japan) equipped with a resonant circuit that selectively amplified the signal at the optical modulation frequency. The stimulated Raman loss signal was then extracted by a lock-in amplifier (HF2LI, Zurich Instrument, Zurich, Switzerland). The output voltage from the lock-in amplifier, which represented the SRS signal, was sampled by a data acquisition card (PCIe-6363, National Instruments, Austin, TX). A LabVIEW program was written for synchronized galvanometer scanning and 2D acquisition of images. SRS imaging was performed by tuning the pump wavelength to 796.8 and 791.8 nm to match the Raman bands of 2850 and 2930 cm^−1^, which correspond to the C–H stretching region of lipid and protein respectively. The excitation power at the sample was ∼40 mW for pump and ∼100 mW for Stokes. Each SRS image contained 400×400 pixels. The pixel dwell time was 10 μs. The field of view of single SRS images was 104×104 μm.

### Single-cell segmentation and quantitative evaluation

Single-cell segmentation was performed using DS-Cellpose, a deep learning-based method, as shown in **Supplementary Fig. 1**. Due to the significant variability in cell shapes, the multi-scale fusion capability of the segmentation network needed to be further enhanced. Based on this idea, a DS module was introduced into the decoder portion of the segmentation network. This module primarily involved adding 3×3 convolutions and up-sampling at each layer of the decoder to obtain segmentation results for that layer. Subsequently, multiple segmentation results with different scale information were further weighted and fused to output the final segmentation result. The DS module improved the multi-scale fusion capability of the segmentation network and the training speed of the model. Training for single-cell segmentation was conducted using 130 SRS images of UEC. The trained model was then used for SRS image segmentation, and the segmented masks were saved. In comparison, single-cell segmentation using Cellpose, StarDist, U-Net, and the watershed method was performed using Python-based tools: Cellpose 2.2.2, Stardist 0.8.3, U-Net, and Fiji (with automatic thresholding for binary mask creation).

Segmentation performance was quantified using three metrics: global IoU, AP_at 0.5_, and cell IoU. The gold standard was determined by human visual inspection.

Global IoU measured the overall overlap between the predicted and ground truth masks and was defined as:

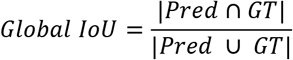

where |·| represented the total area of the cell region.

AP_at 0.5_ assessed over- and under-segmentation and was defined as:

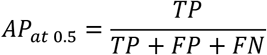

where TP, FP, and FN represent the numbers of true-positive, false-positive, and false-negative segmented cell instances, respectively. A predicted cell was considered a true positive when its intersection-over-union (IoU) with the corresponding ground-truth cell was ≥ 0.5.

Cell IoU quantified the overlap between each segmented cell and its corresponding ground-truth cell and was defined as:

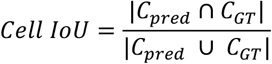

where *C*_pred_ and *C*_GT_ represented the area of each segmented cell and its matched ground-truth cell. This metric provided a more accurate measure of single-cell segmentation performance compared with global IoU, which was less sensitive to under-or over-segmentation.

### Extraction of features from single cells

A total of 20 features were extracted from single-cell segmentation, comprising morphological and compositional features (**Fig. 2a**).

Morphological features included cellular area, eccentricity, equivalent diameter, major axis length, minor axis length, perimeter, shape, nucleus area, cytoplasm area, and the nucleus cytoplasm ratio.

Features such as cellular area, eccentricity, equivalent diameter, major axis length, minor axis length, and perimeter were extracted directly from segmented cell masks using the RegionProps function in the Python Scikit-image library. The major and minor axis lengths corresponded to the principal and minor axes of the ellipse that best fitted the cell contour. The shape was calculated as:

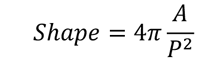

where *A* denoted the cellular area and *P* denoted the perimeter. A perfect circle yielded a value of 1, whereas elongated or indented cells showed smaller values, reflecting a greater perimeter at constant area.

For each field of view in the two-color SRS images, molecular images representing lipid and protein concentrations at each pixel were calculated according to:

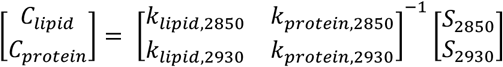

where *k* denoted the contribution of a given molecular component to the SRS signal at a specific Raman shift (derived from the Raman spectra of pure standards), *S* represented the pixel intensity of the SRS image at the corresponding Raman shift, and *C* represented the calculated component concentration. This calculation relied on the linear relationship between SRS intensity and the molar concentration of Raman-active vibrational modes. Single-cell composite images from the lipid (green) and protein (blue) channels were then used to extract compositional features. Each image contained 400 × 400 pixels and was centered on segmented cells.

Based on imaging results, the lipid-channel intensity within nucleus was significantly lower than that of the cytoplasm due to the reduced neutral lipid content in the nucleus. This feature was utilized to separate the nucleus from the cytoplasm in SRS images using grayscale erosion. Following nucleus segmentation in the lipid channel, the cytoplasm area was obtained by subtracting the nucleus area from the total cellular area, and a cytoplasm mask was generated. The nucleus cytoplasm ratio was then calculated as the ratio of nucleus area to cytoplasm area.

Compositional features included LD number, LD distance, LD relative distance, LD area, LD area ratio, lipid intensity, protein intensity, lipid protein ratio, lipid intensity within the cytoplasm, and protein intensity within the cytoplasm.

The YOLOv5 model for LD detection provided five LD related features, including LD number, LD area (approximated from the detection bounding box), ratio of LD area to cellular area (LD area ratio), distance from LD to the centroid (LD distance), and ratio of distance from LD to the centroid and the equivalent diameter of the cell (LD relative distance). The YOLOv5 model processed an image with 400×400 pixels (field of view (FOV) of 104×104 μm^2^) in approximately 5 ms. Training was initialized from the YOLOv5s checkpoint using the default hyperparameters provided in the Ultralytics repository. LDs in approximately 150 SRS images at 2850 cm^-1^, including BCa and non-BCa samples, were manually annotated on the Roboflow platform, and labels were exported in YOLOv5 format. Each image was cropped into 2×2 tiles to facilitate detection of small objects, resized into 640×640 pixels, and augmented via horizontal/vertical flipping, rotation and brightness adjustment (−15 % to +15 %). Background intensity derived from cell-segmentation masks was set to zero before feeding the image into the LD detection model. Retained LDs were required to meet the following criteria: detection confidence > 0.3, and either local contrast > 0.8 or LD area > 10 pixels^2^. Local contrast was calculated from the original image and the image after a morphological closing operation, which replaced local maxima (i.e., LDs) with surrounding lower-intensity pixels. The LD features of each cell were derived from the retained LDs.

Finally, lipid intensity, protein intensity, lipid-to-protein ratio, lipid intensity within the cytoplasm, and protein intensity within the cytoplasm were computed from the molecular images with cell mask and cytoplasm mask.

### PCA and MIL-XGBoosting classification

For each feature combination, the selected single-cell features were transformed using incremental principal component analysis (PCA), yielding 10 principal components (PCs) for subsequent classification. Classification was performed within a multiple instance learning (MIL) framework, in which each participant was treated as a bag containing multiple urinary exfoliated cells as individual instances. XGBoosting was used as the instance-level classifier with 200 estimators, a learning rate of 0.04, a maximum tree depth of 10, and a binary logistic objective. During iterative MIL training, a top-k optimization strategy was applied, in which the k instances with the highest predicted probabilities in misclassified positive bags were assigned positive labels, followed by model retraining until convergence. In this study, k was set to 5. Diagnostic performance was evaluated using leave-one-out cross-validation at the participant level. The probability of each held-out participant was defined as the maximum predicted probability among its constituent cells.

### Large-scale SRS imaging

By implementing an automated wide-field SRS imaging protocol, large-scale imaging was performed on two positive and two negative sections. For each unstained cell smear, 5×10 images were acquired at 10% overlap to generate an FOV of 400×800 μm^2^. After acquisition, a stitching algorithm implemented in MATLAB was used to merge and edge-smooth the images. Subsequently, exfoliated cells were subjected to H&E staining and digitally imaged with a slide scanner. Two certified pathologists independently reviewed the H&E sections and used ImageJ to outline positive cells in red and negative cells in green.

### Statistical analysis

Mann-Whitney U test was applied for group comparisons; and *P* < 0.05 was considered statistically significant. *P* values were calculated with Origin 2025 using Mann-Whitney U test. Differences in LD number between the BCa and non-BCa groups were assessed using a two-tailed unpaired Student’s t-test in GraphPad Prism 10.1.2. Effect size was quantified using eta squared (η^2^), calculated as η^2^ = t^2^ / (t^2^ + df), where t is the t statistic and df is the corresponding degrees of freedom from the same t-test. Classification performance was assessed by ROC analysis; the AUC quantified the model’s ability to discriminate BCa from non-BCa samples. The optimal patient-level classification threshold for each model was determined by maximizing Youden’s index, defined as J = sensitivity+specificity−1, across the candidate thresholds on the ROC curve.

## Data Availability

All data produced in the present study are available upon reasonable request to the authors

https://github.com/jormehao-cyber/DS-Cellpose.git

## Data availability

The primary data supporting the findings are presented within the paper and its Supplementary Information. All other data supporting this study are available from the corresponding author upon reasonable request.

## Code availability

The original DS-Cellpose code is available at https://github.com/jormehao-cyber/DS-Cellpose.git and was adapted to our dataset. MATLAB scripts for tiling and restitching are available upon request. The system-control and data-collection software are proprietary and licensed.

## Funding

This work was supported by National Key R&D Program of China (No. 2025YFF0517900, 2023YFC2415500); PKU-Baidu Fund (No. 2020BD033); Peking University First Hospital Scientific and Technological Achievement Transformation Incubation Guidance Fund (No. 2022CX02); Fundamental Research Funds for the Central Universities (JKF-2025086684620); National Natural Science Foundation of China (No. 82672586).

## Author contributions

X.C., L.Y. and S.Y. conceived and designed the study; Z.H. performed sample preprocessing and SRS imaging; T.F. and J.L. developed the computational methods and analyzed the data with assistance from H.H., X.Z. and D.X.; Y.Z. coordinated clinical sample collection and cytopathological assessment with assistance from S.H. and L.Z.; Z.H., J.L., X.C. and S.Y. drafted and revised the manuscript with input from all authors. All authors reviewed and approved the final manuscript.

## Competing interests

The authors declare no competing commercial or financial interests that could be construed as a potential conflict of interest.

## Notes

### Competing Interest Statement

The authors have declared no competing interest.

### Author Declarations

This study was approved by the Institutional Review Boards of Beijing Cancer Hospital and Beihang University, and written informed consent was obtained from all participants.

## References

1 Sung, H. et al. Global cancer statistics 2024: GLOBOCAN estimates of incidence and mortality worldwide for 34 cancers in 186 countries. CA: a cancer journal for clinicians 76, e70090 (2026). 10.3322/caac.70090

2 van der Heijden, A. G. et al. European Association of Urology Guidelines on Muscle-invasive and Metastatic Bladder Cancer: Summary of the 2025 Guidelines. European urology 87, 582–600 (2025). 10.1016/j.eururo.2025.02.019

3 Tran, L., Xiao, J.-F., Agarwal, N., Duex, J. E. & Theodorescu, D. Advances in bladder cancer biology and therapy. Nature Reviews Cancer 21, 104–121 (2021). 10.1038/s41568-020-00313-1

4 Kobayashi, T., Owczarek, T. B., McKiernan, J. M. & Abate-Shen, C. Modelling bladder cancer in mice: opportunities and challenges. Nature Reviews Cancer 15, 42–54 (2015). 10.1038/nrc3858

5 Witjes, J. A. et al. EAU-ESMO Consensus Statements on the Management of Advanced and Variant Bladder Cancer—An International Collaborative Multistakeholder Effort†: Under the Auspices of the EAU-ESMO Guidelines Committees. European urology 77, 223–250 (2020). 10.1016/j.eururo.2019.09.035

6 Flaig, T. W. et al. NCCN Guidelines® Insights: Bladder Cancer, Version 2.2022. Journal of the National Comprehensive Cancer Network : JNCCN 20, 866–878 (2022). 10.6004/jnccn.2022.0041

7 Babjuk, M. et al. European Association of Urology Guidelines on Non-muscle-invasive Bladder Cancer (TaT1 and Carcinoma In Situ) - 2019 Update. European urology 76, 639–657 (2019). 10.1016/j.eururo.2019.08.016

8 Wang, Z. et al. Evaluation of the NMP22 BladderChek test for detecting bladder cancer: a systematic review and meta-analysis. Oncotarget 8, 100648–100656 (2017). 10.18632/oncotarget.22065

9 Siguencia, F. et al. Diagnostic Performance of Urinary Biomarker Tests in Detecting Bladder Cancer: A Systematic Review, Meta-analysis, and Network Meta-analysis. Urology 210, 145–152 (2026). 10.1016/j.urology.2026.01.035

10 Tomiyama, E., Fujita, K., Hashimoto, M., Uemura, H. & Nonomura, N. Urinary markers for bladder cancer diagnosis: A review of current status and future challenges. International Journal of Urology 31, 208–219 (2024). 10.1111/iju.15338

11 Chen, X. et al. Urine DNA methylation assay enables early detection and recurrence monitoring for bladder cancer. The Journal of clinical investigation 130, 6278–6289 (2020). 10.1172/jci139597

12 Keum, C. et al. Diagnosis of early-stage bladder cancer via unprocessed urine samples at the point of care. Nature biomedical engineering 9, 1026–1038 (2025). 10.1038/s41551-024-01298-0

13 Grausenburger, R., Herek, P., Shariat, S. F. & Englinger, B. Recent contributions of single-cell and spatial profiling to the understanding of bladder cancer. Current opinion in urology 34, 236–243 (2024). 10.1097/mou.0000000000001183

14 Lindskrog, S. V. et al. Single-nucleus and Spatially Resolved Intratumor Subtype Heterogeneity in Bladder Cancer. European urology open science 51, 78–88 (2023). 10.1016/j.euros.2023.03.006

15 Pavlova, N. N., Zhu, J. & Thompson, C. B. The hallmarks of cancer metabolism: Still emerging. Cell metabolism 34, 355–377 (2022). 10.1016/j.cmet.2022.01.007

16 Vogel, F. C. E., Chaves-Filho, A. B. & Schulze, A. Lipids as mediators of cancer progression and metastasis. Nature Cancer 5, 16–29 (2024). 10.1038/s43018-023-00702-z

17 Hoy, A. J., Nagarajan, S. R. & Butler, L. M. Tumour fatty acid metabolism in the context of therapy resistance and obesity. Nature Reviews Cancer 21, 753–766 (2021). 10.1038/s41568-021-00388-4

18 Jin, Y., Tan, Y., Wu, J. & Ren, Z. Lipid droplets: a cellular organelle vital in cancer cells. Cell Death Discovery 9, 254 (2023). 10.1038/s41420-023-01493-z

19 Xiong, K. et al. RNF112 Facilitates Ubiquitin-Mediated Degradation of c-Myc, Suppressing Proliferation, Migration and Lipid Synthesis in Bladder Cancer. Advanced science (Weinheim, Baden-Wurttemberg, Germany) 12, e2408311 (2025). 10.1002/advs.202408311

20 Wang, X. et al. PIN1 Prolyl Isomerase Promotes Initiation and Progression of Bladder Cancer through the SREBP2-Mediated Cholesterol Biosynthesis Pathway. Cancer Discov 15, 633–655 (2025). 10.1158/2159-8290.Cd-24-0866

21 Tao, T. et al. Down-regulation of PKM2 decreases FASN expression in bladder cancer cells through AKT/mTOR/SREBP-1c axis. Journal of Cellular Physiology 234, 3088–3104 (2019). 10.1002/jcp.27129

22 Deng, J. et al. Metformin targets Clusterin to control lipogenesis and inhibit the growth of bladder cancer cells through SREBP-1c/FASN axis. Signal Transduction and Targeted Therapy 6, 98 (2021). 10.1038/s41392-021-00493-8

23 Liu, P. et al. m(6)A-induced lncDBET promotes the malignant progression of bladder cancer through FABP5-mediated lipid metabolism. Theranostics 12, 6291–6307 (2022). 10.7150/thno.71456

24 Freudiger, C. W. et al. Label-free biomedical imaging with high sensitivity by stimulated Raman scattering microscopy. Science (New York, N.Y.) 322, 1857–1861 (2008). 10.1126/science.1165758

25 Min, W.Cheng, J.-X. & Ozeki, Y. Theory, innovations and applications of stimulated Raman scattering microscopy. Nature Photonics 19, 803–816 (2025). 10.1038/s41566-025-01707-z

26 Cheng, J.-X. et al. Advanced vibrational microscopes for life science. Nature methods 22, 912–927 (2025). 10.1038/s41592-025-02655-w

27 Li, Y., Liu, J. & Yue, S. Stimulated Raman Scattering Imaging Enabled Biomarker Discovery for Precision Medicine. Chemical & Biomedical Imaging 3, 805 – 827 (2025).

28 Jia, H. & Yue, S. Stimulated Raman Scattering Imaging Sheds New Light on Lipid Droplet Biology. The Journal of Physical Chemistry B 127, 2381–2394 (2023). 10.1021/acs.jpcb.3c00038

29 Zhang, S., He, Y. & Yue, S. Coherent Raman scattering imaging of lipid metabolism in cancer. Journal of Innovative Optical Health Sciences 16, 2230015 (2023). 10.1142/s1793545822300154

30 Chen, T., Yavuz, A. & Wang, M. C. Dissecting lipid droplet biology with coherent Raman scattering microscopy. Journal of Cell Science 135 (2021). 10.1242/jcs.252353

31 Liu, J., Liu, R., Zhang, X., Chen, X. & Yue, S. Deep Learning-Assisted Coherent Raman Scattering Microscopy. Advanced Intelligent Discovery n/a, 202500072 (2025). 10.1002/aidi.202500072

32 Kondepudi, A. et al. Foundation models for fast, label-free detection of glioma infiltration. Nature 637, 439–445 (2025). 10.1038/s41586-024-08169-3

33 Liu, Z. et al. Ultrarapid deep 3D histology enables intraoperative mapping of glioma infiltration. Cell 10.1016/j.cell.2026.07.026

34 Chen, X. et al. Accurate and Rapid Detection of Peritoneal Metastasis from Gastric Cancer by AI-Assisted Stimulated Raman Molecular Cytology. Advanced science (Weinheim, Baden-Wurttemberg, Germany) 10, e2300961 (2023). 10.1002/advs.202300961

35 Fang, T. et al. MICLEAR: Intelligent Molecular Cytology for Intraoperative Margin Assessment of Pancreatic Ductal Adenocarcinoma. Medical Image Analysis, 104181 (2026). 10.1016/j.media.2026.104181

36 Filho, A. M., Briganti, A., Jemal, A. & Bray, F. Bladder Cancer Incidence and Mortality: A Global Overview and Recent Trends. European urology 89, 426–436 (2026). 10.1016/j.eururo.2025.12.011

37 Lee, P. J., Owens, C. L., Lithgow, M. Y., Jiang, Z. & Fischer, A. H. Causes of false-negative for high-grade urothelial carcinoma in urine cytology. Diagnostic Cytopathology 44, 994–999 (2016). 10.1002/dc.23621

38 Velez Torres, J. M. et al. Urine Cytology Findings in Cases of Pseudocarcinomatous Urothelial Hyperplasia of the Bladder Often Represent a Diagnostic Challenge. Archives of Pathology & Laboratory Medicine 147, 716–721 (2023). 10.5858/arpa.2022-0125-OA

39 Lin, T. et al. Prospective evaluation of fluorescence in situ hybridization for diagnosing urothelial carcinoma. Oncology letters 13, 3928–3934 (2017). 10.3892/ol.2017.5926

40 Dimashkieh, H. et al. Evaluation of urovysion and cytology for bladder cancer detection: a study of 1835 paired urine samples with clinical and histologic correlation. Cancer cytopathology 121, 591–597 (2013). 10.1002/cncy.21327

41 Barkan, G. A. et al. The Paris System for Reporting Urinary Cytology: The Quest to Develop a Standardized Terminology. Advances in anatomic pathology 23, 193–201 (2016). 10.1097/pap.0000000000000118

42 Planz, B. et al. The role of urinary cytology for detection of bladder cancer. European journal of surgical oncology : the journal of the European Society of Surgical Oncology and the British Association of Surgical Oncology 31, 304–308 (2005). 10.1016/j.ejso.2004.12.008

43 Becker, G. J., Garigali, G. & Fogazzi, G. B. Advances in Urine Microscopy. Am J Kidney Dis 67, 954–964 (2016). 10.1053/j.ajkd.2015.11.011

44 Haq, K. & Patel, D. M. Urinalysis: Interpretation and Clinical Correlations. Med Clin North Am 107, 659–679 (2023). 10.1016/j.mcna.2023.03.002

45 Yosef, H. K. et al. Noninvasive Diagnosis of High-Grade Urothelial Carcinoma in Urine by Raman Spectral Imaging. Anal Chem 89, 6893–6899 (2017). 10.1021/acs.analchem.7b01403

46 Stringer, C., Wang, T., Michaelos, M. & Pachitariu, M. Cellpose: a generalist algorithm for cellular segmentation. Nature methods 18, 100–106 (2021). 10.1038/s41592-020-01018-x

47 Jocher, G. YOLOv5 by Ultralytics (Version 7.0). <https://github.com/ultralytics/yolov5> (2020).

48 Tao, L. et al. FTO modifies the m6A level of MALAT and promotes bladder cancer progression. Clin Transl Med 11, e310 (2021). 10.1002/ctm2.310

49 Chen, X., Zhang, Z., Liao, W. & Zhao, Y. Assessment tool based on fatty acid metabolic signatures for predicting the prognosis and treatment response in bladder cancer. Heliyon 9, e22768 (2023). 10.1016/j.heliyon.2023.e22768

50 Moreira, I. B. et al. Neolactotetraosylceramide enables urinary detection of bladder cancer. Cell Rep Med 6, 102246 (2025). 10.1016/j.xcrm.2025.102246

51 Zeng, Y. et al. m6A-Mediated Induction of 7-Dehydrocholesterol Reductase Stimulates Cholesterol Synthesis and cAMP Signaling to Promote Bladder Cancer Metastasis. Cancer Res 84, 3402–3418 (2024). 10.1158/0008-5472.Can-23-3703

