## Supplementary Fig. 1-17; Supplementary Table 1-4. for "LUCID: Lipid-featured Urine Cytology with Intelligent Detection for Non-Invasive Early Diagnosis of Bladder Cancer"

[1]National Medical Innovation Platform for Industry-Education Integration in Advanced Medical Devices (Interdiscipline of Medicine and Engineering); Key Laboratory of Innovation and Transformation of Advanced Medical Devices, Ministry of Industry and Information Technology; Institute of Medical Photonics, Beijing Advanced Innovation Center for Biomedical Engineering; School of Biological Science and Medical Engineering, and with the School of Engineering Medicine, Beihang University, Beijing 100191, China

[2]Department of Urology, Peking University First Hospital, Beijing, China; Institute of Urology, Peking University, Beijing 100034, China

[3]Optical Bioimaging Laboratory, Department of Biomedical Engineering, College of Design and Engineering, National University of Singapore, Singapore 117576

[#]These authors contributed equally.

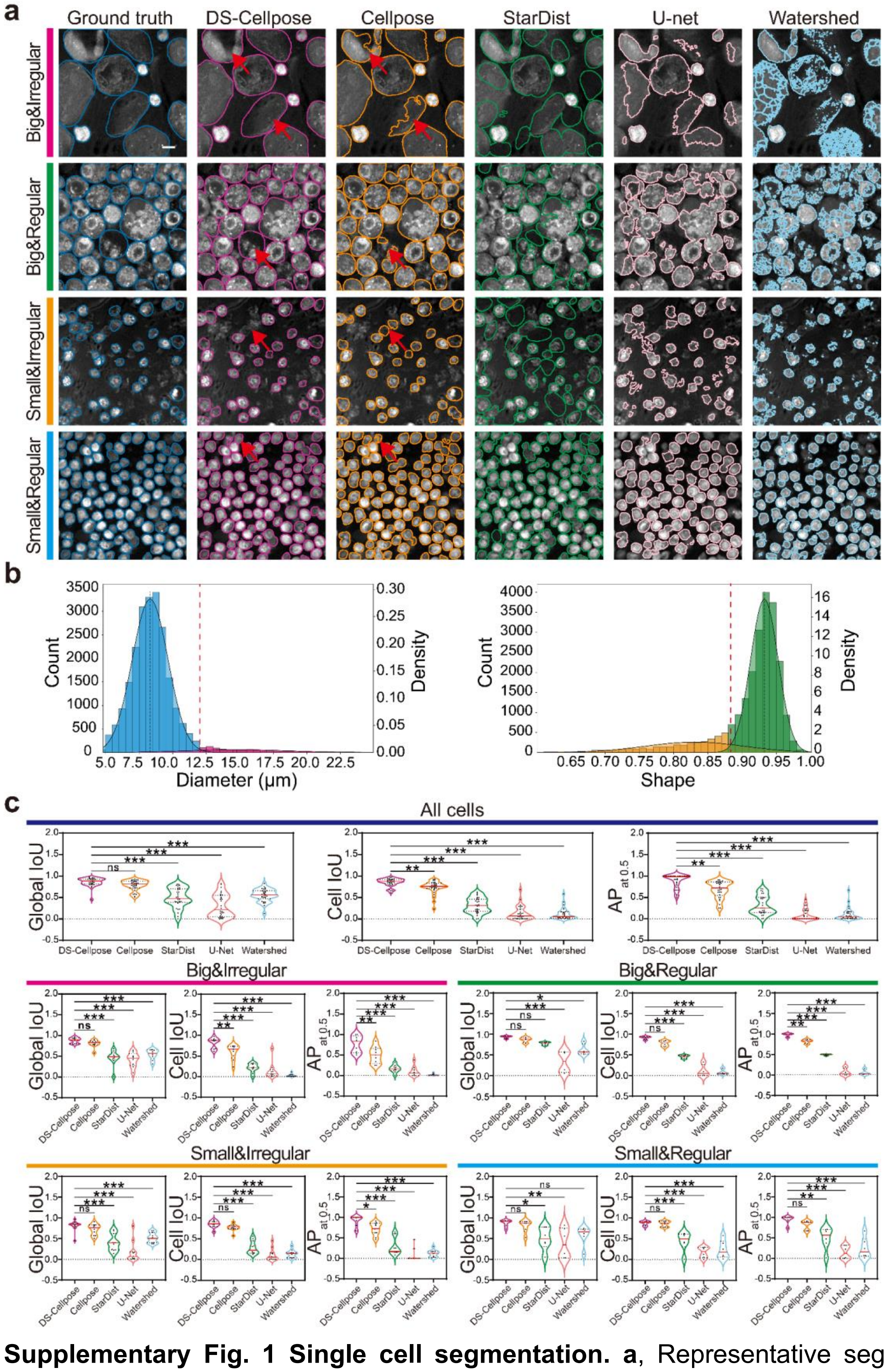


**Supplementary Fig. 1 Single cell segmentation. a**, Representative segmentation results for cells with different shape and size, by using DS-cellpose, Cellpose, StarDist, U-net, and Watershed models. Scale bar: 10μm. **b,** Histograms showing the distributions of cell diameter and shape in **a**. Cells were classified according to diameter and shape using Gaussian mixture models (GMMs), resulting in large and small cell groups and regular and irregular shape groups, respectively. Dashed red lines indicate the classification thresholds. **c**, Evaluation of the segmentation performance, measured by global IoU, cell IoU and $AP_{at\ 0.5}$. Violin plots (mean: red horizontal line, quartiles: black horizontal dashed line; ns: non-significant, *: $p$ <0.05, **: $p$ <0.01, ***: $p$ <0.001, ****: p <0.0001, Mann-Whitney U test).

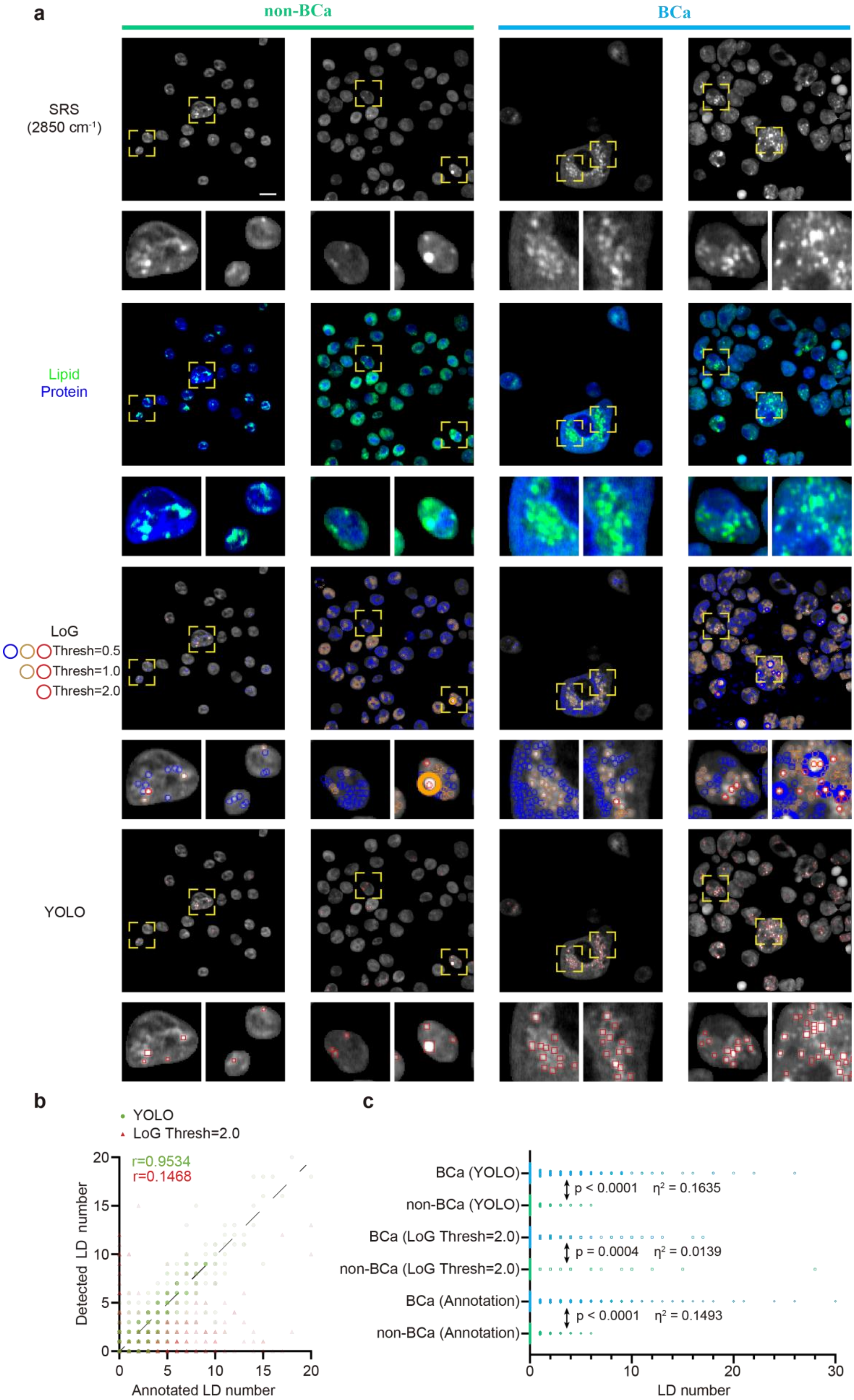

a
non-BCa
BCa
SRS
(2850 cm-1)
Lipid
Protein
LoG
Thresh=0.5
Thresh=1.0
Thresh=2.0
YOLO
b
YOLO
LoG Thresh=2.0
r=0.9534
r=0.1468
Detected LD number
Annotated LD number
c
BCa (YOLO)
non-BCa (YOLO)
BCa (LoG Thresh=2.0)
non-BCa (LoG Thresh=2.0)
BCa (Annotation)
non-BCa (Annotation)
p < 0.0001 η2 = 0.1635
p = 0.0004 η2 = 0.0139
p < 0.0001 η2 = 0.1493
LD number

**Supplementary Fig. 2 Deep learning assisted LD detection.** **a**, Representative raw SRS images, corresponding two-color images (Lipid: Green color; Protein: Blue color) and results of LD detection by LoG (red/orange/blue circle) and YOLO (red contour). The detection with LoG used a series of threshold parameter values. To facilitate visualization, zoom-in images of the representative areas (as indicated by yellow boxes) are shown. **b**, Number of detected LDs against annotated ones. *r* denotes Pearson correlative coefficient. **c**, Number of detected LDs in BCa and non-BCa samples. P-values denote T test's statistical significance, while $\eta^2$ (eta-squared) indicates the effect size. Totally 3194 cells from five BCa and non-BCa samples were included in **b** and **c**.

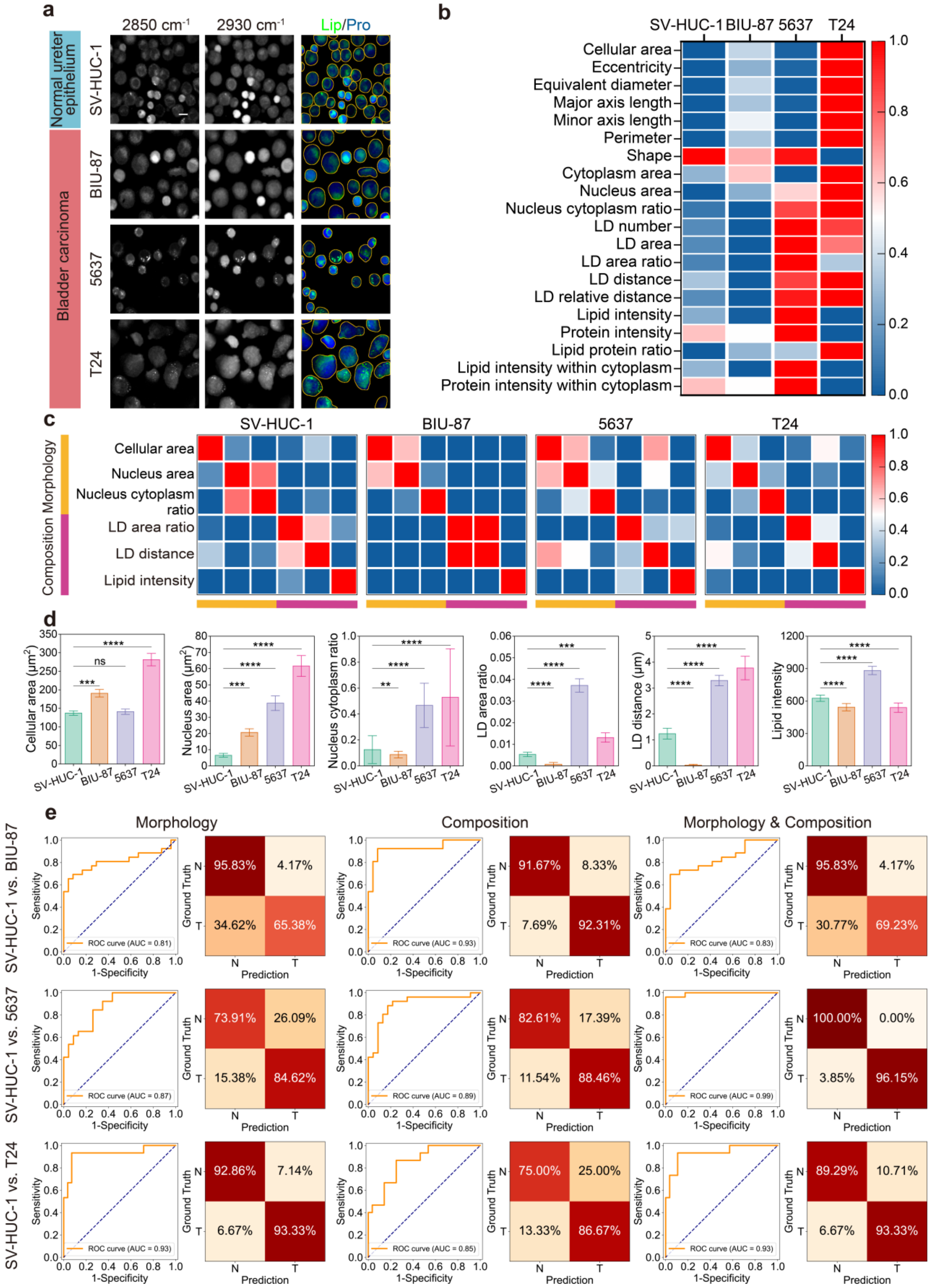

**Supplementary Fig. 3 Differential single-cell features between BCa and normal cell lines. a**, Representative two-color SRS images of normal bladder epithelium (SV-

HUC-1), bladder carcinoma cells (BIU-87, 5637, T24). Lipid channel: Green color; Protein channel: Blue color. Single cell segmentation: Yellow contour. Scale bar: 10μm. **b**, Single-cell feature map of the normal and BCa cell lines. **c**, Correlation coefficient mapping of three morphological features and three compositional features of the normal and BCa cell lines. **d**, The bar chart of the differential features in morphology and composition of the cell lines. The top of each box represents the cell-level mean, and the error bars denote the standard error (SE) of the data. Statistical significance is indicated by the p value from the Mann-Whitney U test (ns: non-significant, *: $p < 0.05$, **: $p < 0.01$, ***: $p < 0.001$, ****: $p < 0.0001$, n: SV-HUC-1 = 122, BIU-87 = 126, 5637 = 119, T24 = 89). **e**, Receiver Operating Characteristic (ROC) curve and confusion matrix for BCa cell detection based on morphology and/or composition features, by Logistic regression (LR) model. AUC: area under the ROC curve.

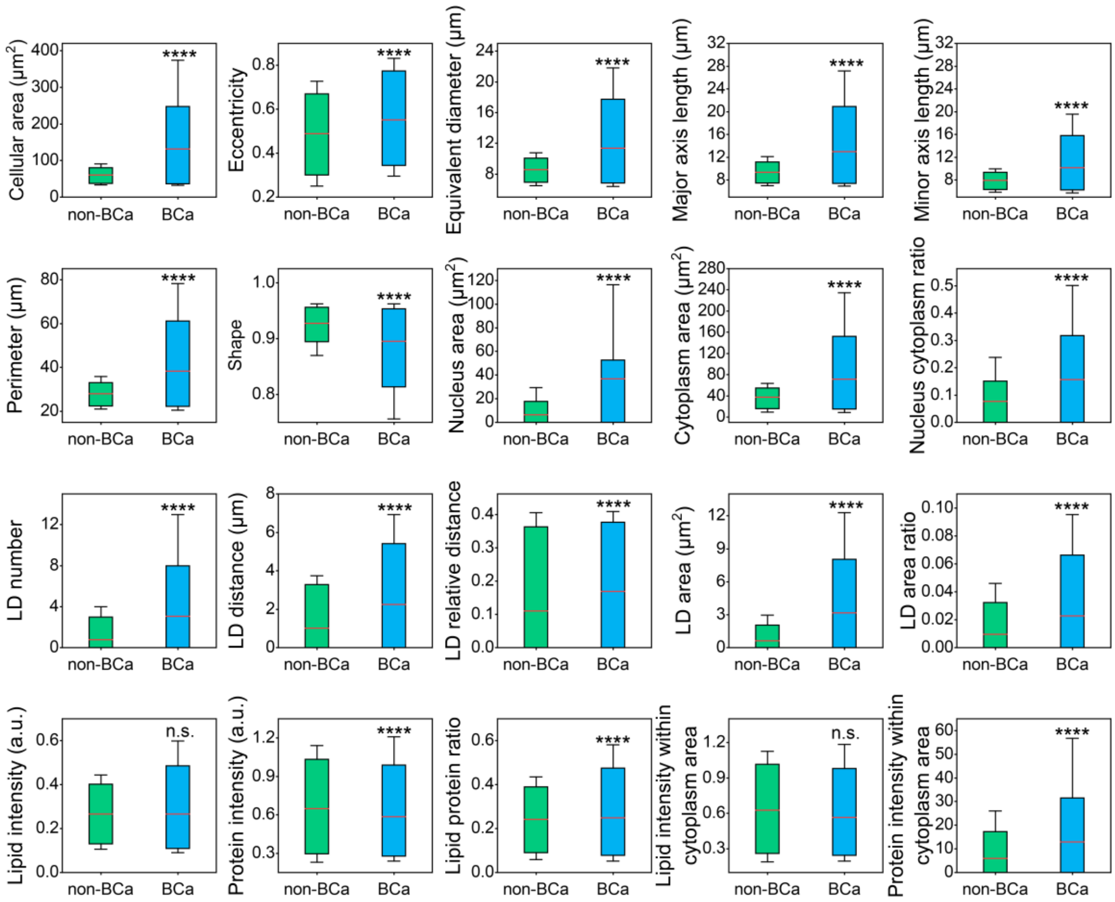


**Supplementary Fig. 4 Box plots of 20 features of single cells in non-BCa and BCa cases.** The red lines in the box indicate the average value (ns: *P* >0.05, *: *P* <0.05, **: *P* <0.01, ***: *P* <0.001, ****: *P* <0.0001, Mann-Whitney U test).

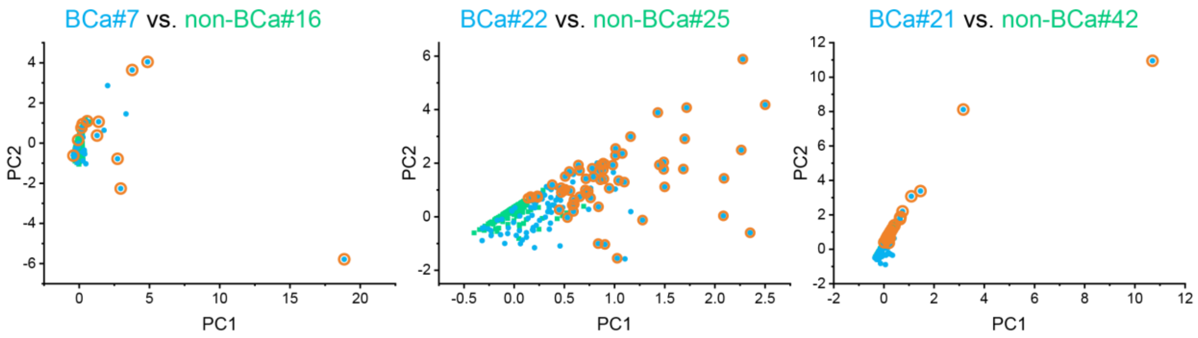


**Supplementary Fig. 5 The complete data of marker cell identification by PCA-MIL algorithm.** PC1 versus PC2 for BCa#7 and non-BCa#16, BCa#22 and non-BCa#25, and BCa#21 and non-BCa#42. Highlighted points denote significant marker cells.

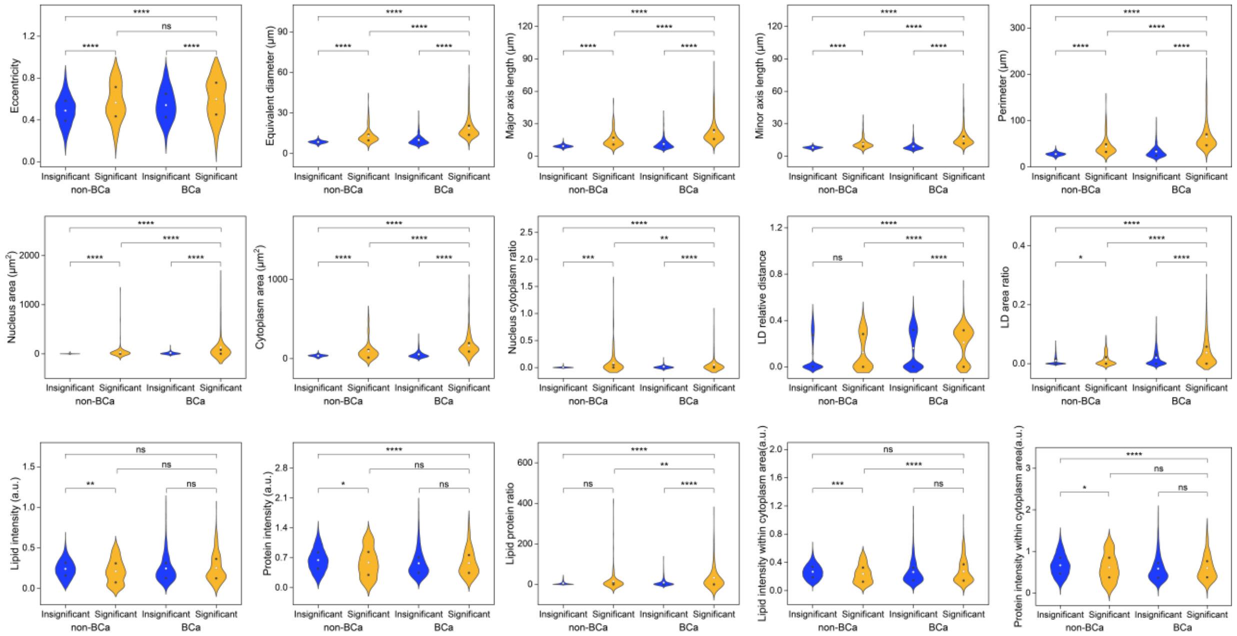

**Supplementary Fig. 6 Comparison of marker cell features at cell-level.** Violin plot shown the distribution of 15 features in single marker cells from non-BCa and BCa samples (ns: $P$ >0.05, *: $P$ <0.05, **: $P$ <0.01, ***: $P$ <0.001, ****: $P$ <0.0001, Mann-Whitney U test). The open circles within the boxes represent the mean, while the black dots indicate the 25% and 75% percentiles.

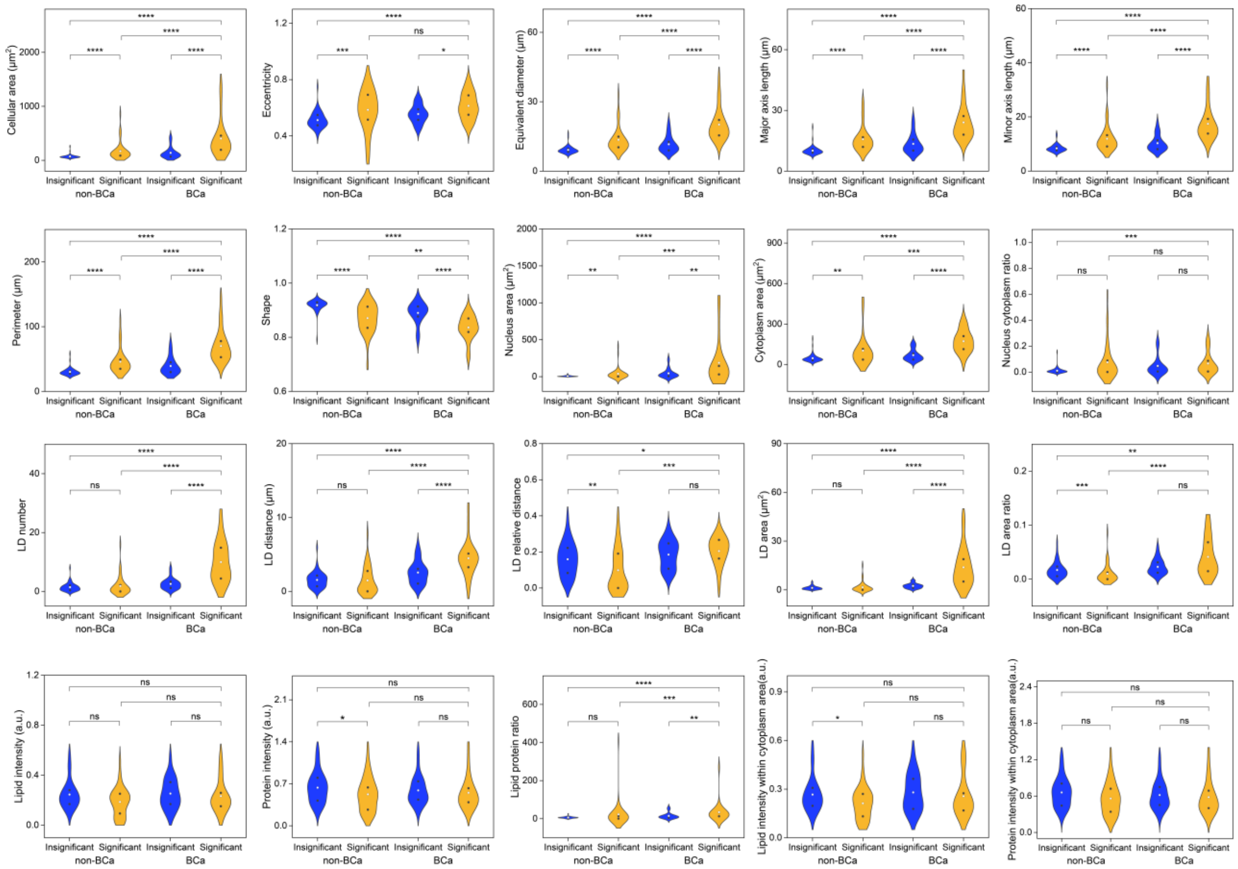


**Supplementary Fig. 7 Comparison of marker cell features at patient-level.** Violin plot showing 20 features of the patient-level from non-BCa and BCa samples (ns: *P* >0.05, *: *P* <0.05, **: *P* <0.01, ***: *P* <0.001, ****: *P* <0.0001, Mann-Whitney U test). The open circles within the boxes represent the mean, while the black dots indicate the 25% and 75% percentiles.

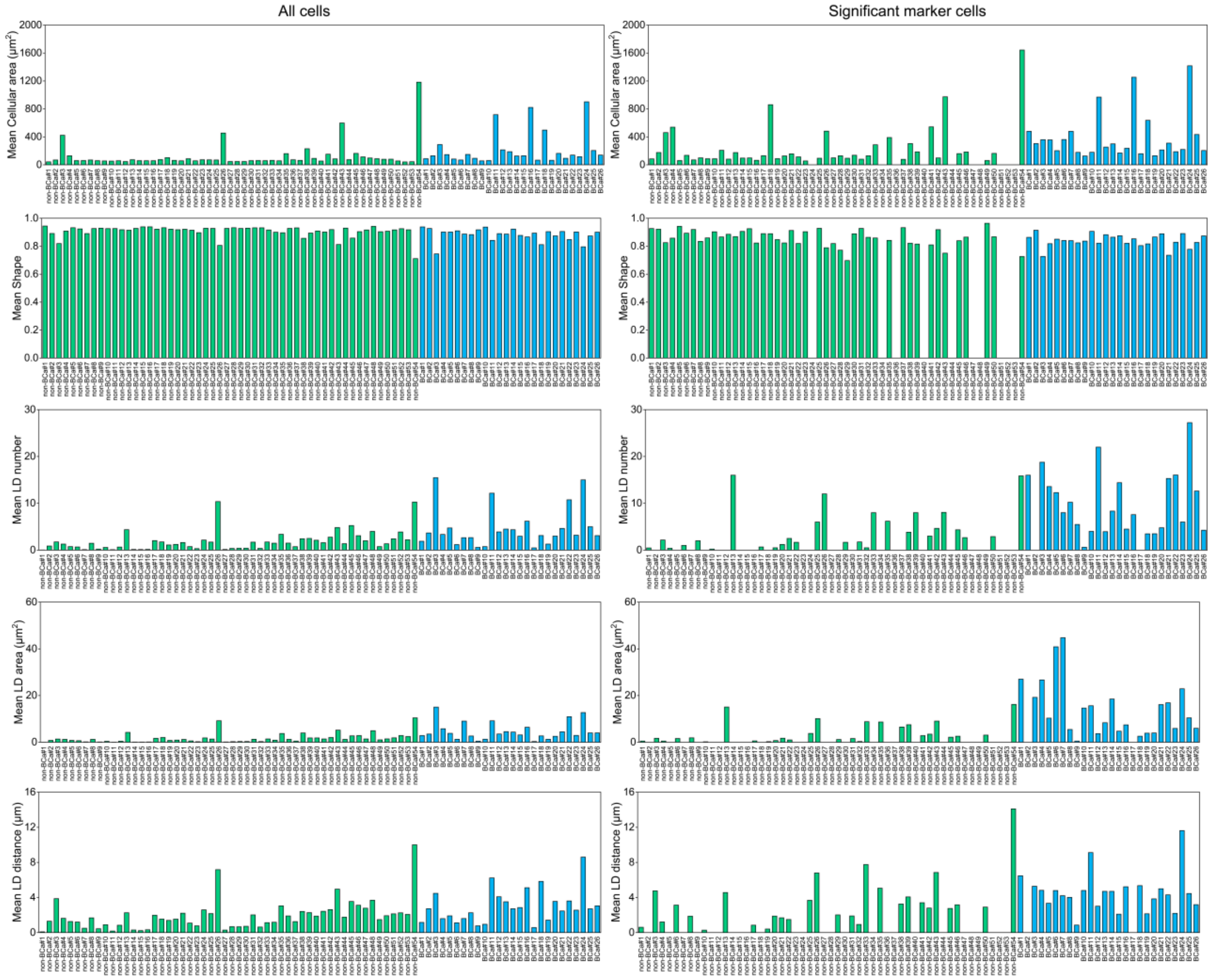


**Supplementary Fig. 8 Comparison of feature values among all cells and significant marker cells of 80 patients.** The LD feature values are averaged by the cell number of each patient. All cells are included to calculate mean feature values.

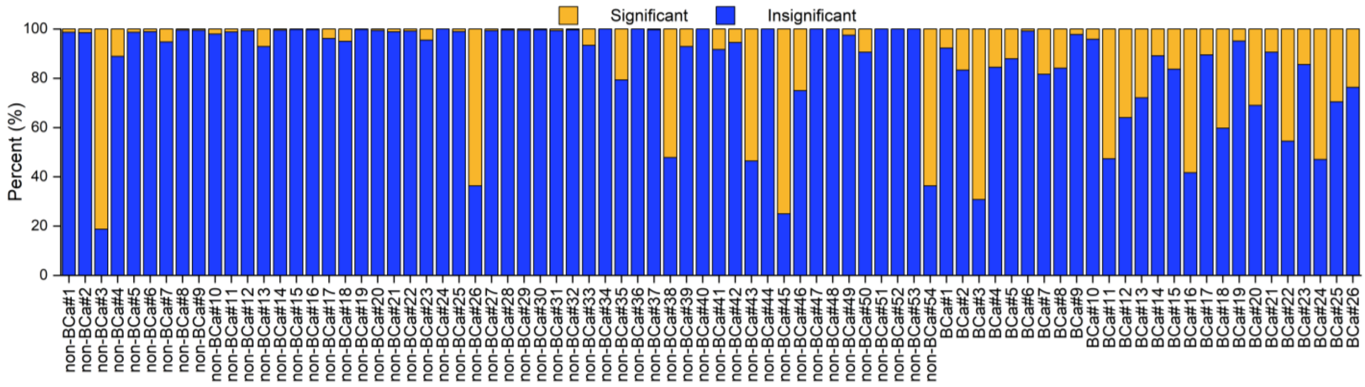


**Supplementary Fig. 9 Percentage histogram of significant vs. insignificant marker cell across all cases.**

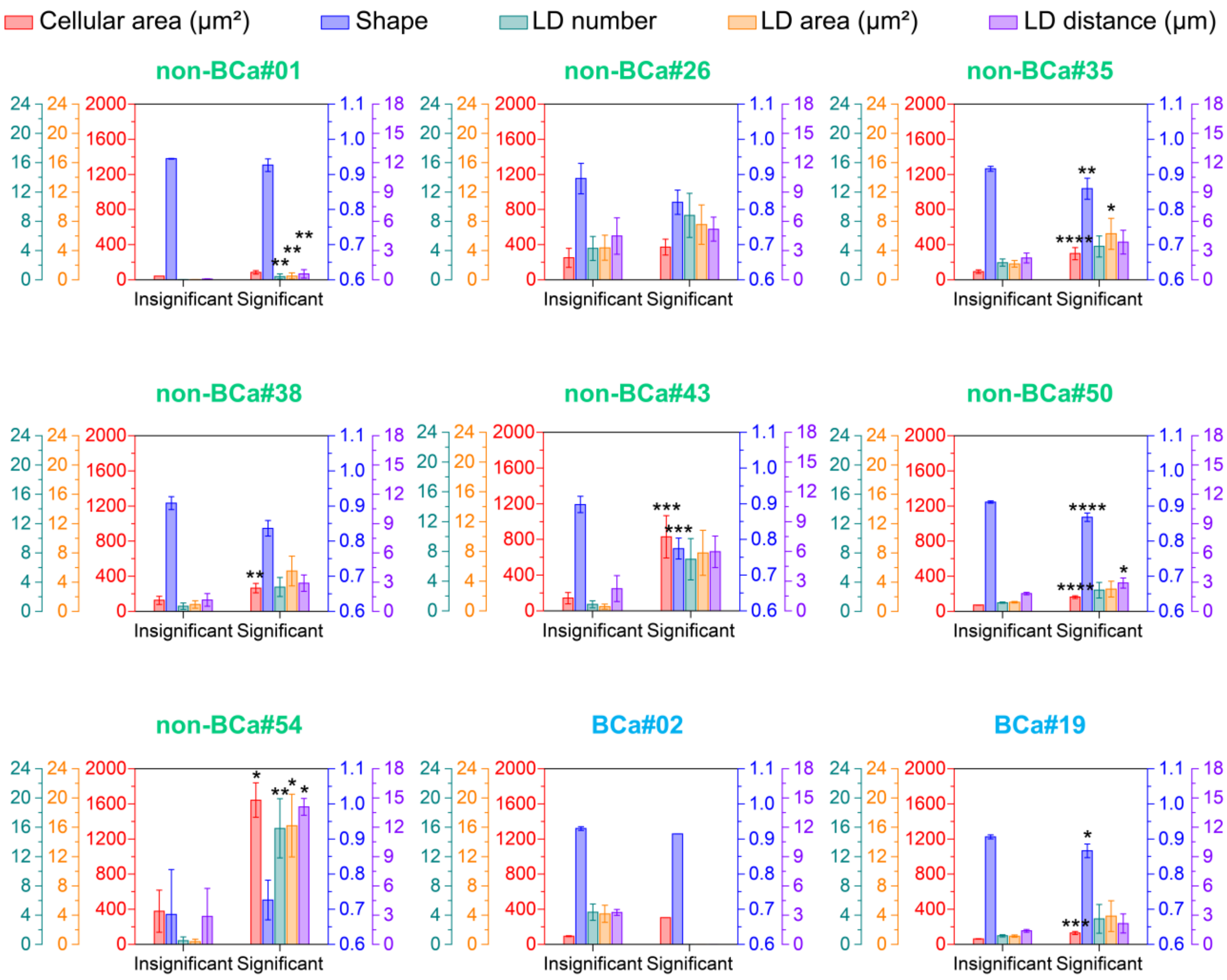


**Supplementary Fig. 10 Features statistics of nine samples failed in LUCID prediction.** Quantitative comparison of representative cellular features between significant and insignificant marker cells in two BCa and seven non-BCa samples misclassified by LUCID. Bars represent group means, and error bars indicate the standard deviation (SD). Statistical significance was assessed using the Mann–Whitney U test (ns: $P$ >0.05, *: $P$ <0.05, **: $P$ <0.01, ***: $P$ <0.001, ****: $P$ <0.0001).

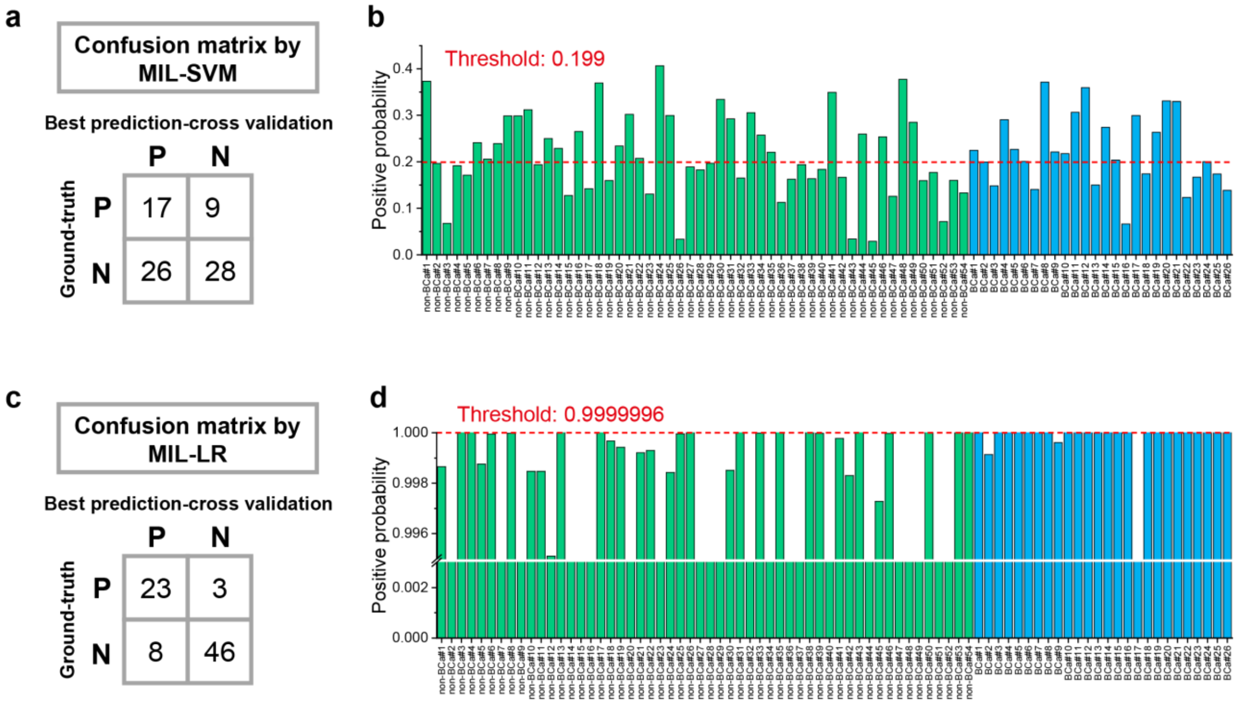


**Supplementary Fig. 11 BC detection with PCA-MIL based on machine learning models other than XGBoosting. a**, the confusion matrix and **b**, the predicted positive probabilities of PCA-MIL based on SVM. **c**, the confusion matrix and **d**, the predicted positive probabilities of PCA-MIL based on LR.

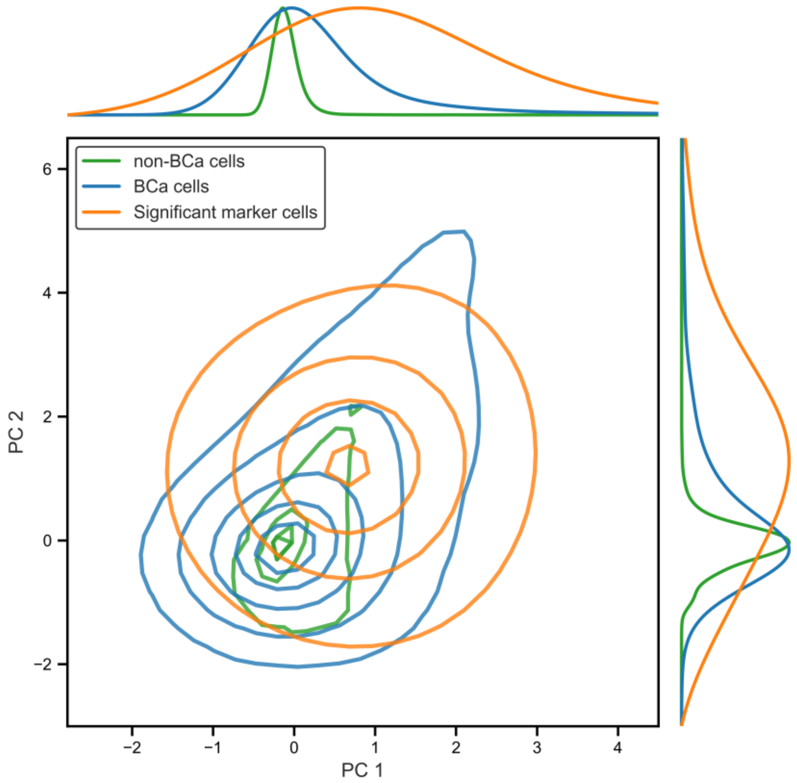


**Supplementary Fig. 12 Visualization of the single-cell feature space via PCA**. The bivariate kernel density estimation (KDE) plot illustrates the distribution of cells in the space of the first two principal components (PC1 and PC2). Green: cells from non-BCa samples, Blue: cells from BCa samples, Orange: significant marker cells. The marginal plots (top and right) show the corresponding univariate density estimates (with Min-Max normalization), highlighting the sharp peaks of non-BCa and BCa cells versus the flattened, broad curves and higher PC1/PC2 values of significant marker cells.

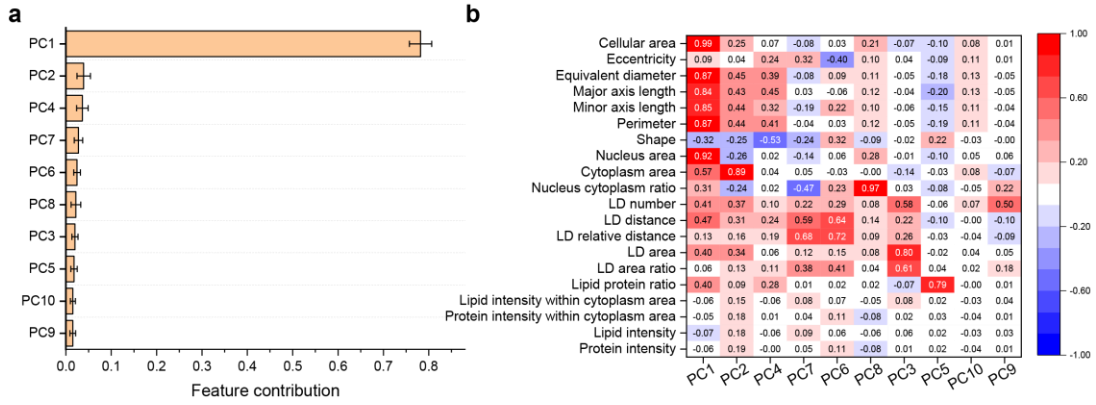


**Supplementary Fig. 13 Characterization of the morphological and lipid-related feature space via PCA and XGBoost feature importance analysis. a,** Bar plot showing the XGBoost-derived feature importance scores, calculated as the average scores across models from the leave-one-out cross-validation procedure. Error bars represent the variation across the validation rounds, revealing the overwhelming predominance of PC1 compared to the other principal components in the classification model. **b,** Heatmap depicting the correlation matrix between specific morphological parameters, lipid-related features, and the first ten PCs (PC1–PC10). The color scale indicates the correlation coefficients, ranging from blue (negative correlation, -1.00) to red (positive correlation, 1.00).

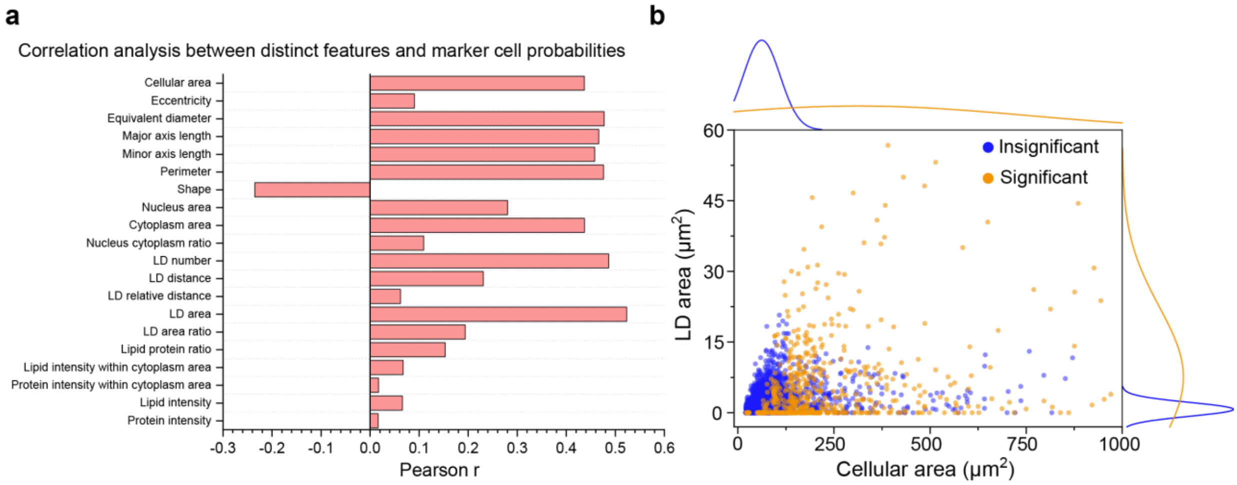


**Supplementary Fig. 14 Pearson correlation analysis between distinct cellular features and marker cell probabilities. a,** Bar plot displaying the Pearson correlation coefficients (Pearson r) between cellular features and the probability of being a significant marker cell. **b,** Scatter plot illustrating the distribution of individual cells based on cellular area and LD area, with marginal univariate density estimates shown on the top and right axes. Blue: insignificantly relevant cells, Yellow: significant marker cells.

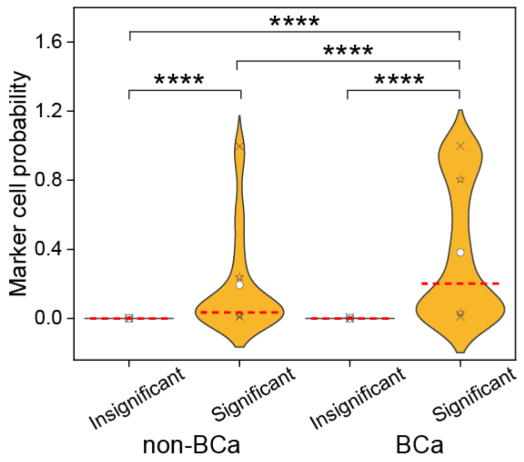

**Supplementary Fig. 15** Violin plots illustrating the distribution of marker cell probability among significant and insignificant marker cells across all non-BCa and BCa samples. Statistical significance between groups was determined using the Mann-Whitney U test (ns: $P > 0.05$, *: $P < 0.05$, **: $P < 0.01$, ***: $P < 0.001$, ****: $P < 0.0001$). Within each violin plot, the symbols and lines denote the following statistical parameters: ×: 1st and 99th percentiles; ☆: 25th and 75th percentiles; ○: mean value; Red dashed lines: median.

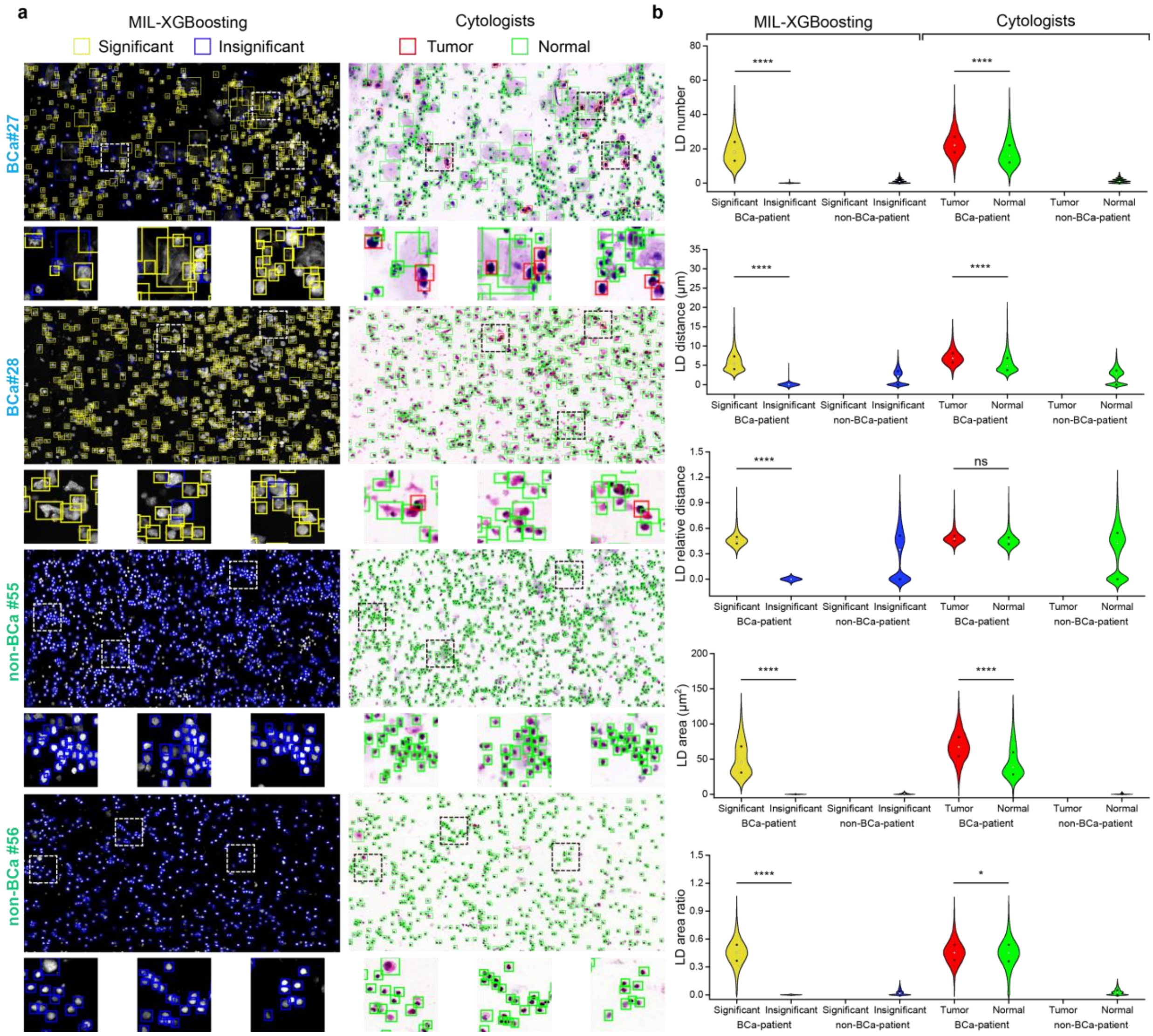


**Supplementary Fig. 16 Marker cell identification and cytologists annotations. a**, Significant marker cells identified by MIL-XGBoosting algorithm and cell-wise annotations in four patients with H&E cytological images (BCa#27, BCa#28, non-BCa#55, and non-BCa#56). Scale bars, 100 μm. **b**, Quantitative comparison between LD features of marker or annotated cells in positive and negative samples (ns: P >0.05, *: P <0.05, **: P <0.01, ***: P <0.001, ****: P <0.0001, Mann-Whitney U test). Percentiles of 25%, and 75% are labelled in the violin plot from bottom to top. The violin plot is plotted to have the same area.

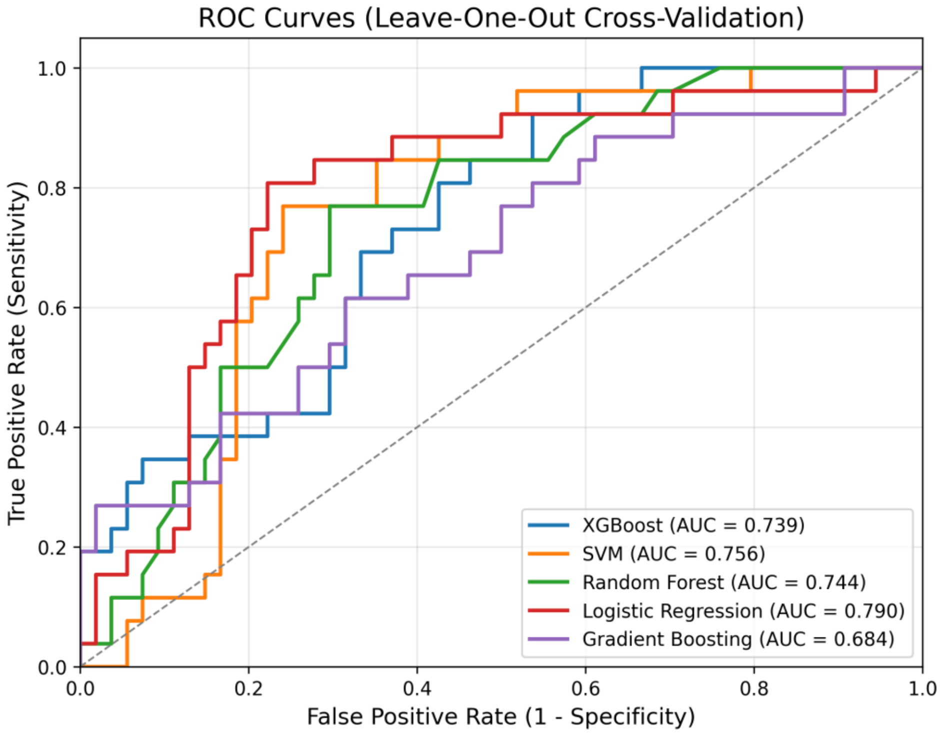


**Supplementary Fig. 17. ROC curves of multiple classifiers using an average-pooling feature strategy.** The classification was performed within a Leave-One-Out Cross-Validation framework. The average pooling scheme aggregated the cellular features of each patient into a single feature vector by calculating their mean. The plot displays the ROC curves for XGBoost, SVM, Random Forest, Logistic Regression, and Gradient Boosting, with corresponding AUC values of 0.739, 0.756, 0.744, 0.790, and 0.684, respectively.

**Supplementary Table 1. Clinical information of normal donors and patients**

| | Non-BCa cohort<br>n=54 | BCa cohort<br>n=26 |
|---|---|---|
| **Age** | | |
| <60 | 13 (24.1%) | 5 (19.2%) |
| ≥60 | 41 (75.9%) | 21 (80.8%) |
| Mean age (range) | 62.7 (20-87) | 67.5 (41-83) |
| **Sex** | | |
| Male | 46 (85.2%) | 18 (69.2%) |
| Female | 8 (14.8%) | 8 (30.8%) |
| **non-BCa** | | |
| Normal | 6 (11.1%) | N/A |
| Urinary stones | 4 (7.4%) | N/A |
| Interstitial cystitis | 3 (5.6%) | N/A |
| Urinary incontinence | 4 (7.4%) | N/A |
| BPH | 37 (68.5%) | N/A |
| **Tumor stage** | | |
| Ta | N/A | 5 (19.2%) |
| T1 | N/A | 17 (65.4%) |
| T2 | N/A | 3 (11.5%) |
| T3 | N/A | 1 (3.8%) |
| **Tumor grade** | | |
| LG | N/A | 10 (38.5%) |

| | | |
|---|---|---|
| HG | N/A | 16 (61.5%) |
| **Tumor size** | | |
| ≤ 1.5 cm | N/A | 8 (30.8%) |
| 1.5-3 cm | N/A | 8 (30.8%) |
| ≥ 3 cm | N/A | 10 (38.4%) |
| **Number of tumor** | | |
| Single | N/A | 4 (15.4%) |
| Multiple | N/A | 22 (84.6%) |

**Supplementary Table 2. Mann-Whitney U test *P* value of 20 features between BCa (n = 26) and non-BCa (n = 54) samples.**

| Feature ID | Features | *P* value | Asterisk |
|---|---|---|---|
| 1 | Cellular area | 3.89E-05 | **** |
| 2 | Eccentricity | 3.14E-02 | * |
| 3 | Equivalent diameter | 1.74E-04 | *** |
| 4 | Major axis length | 9.69E-05 | **** |
| 5 | Minor axis length | 4.52E-04 | *** |
| 6 | Perimeter | 1.30E-04 | *** |
| 7 | Shape | 3.30E-04 | *** |
| 8 | Nucleus area | 2.71E-04 | *** |
| 9 | Cytoplasm area | 8.90E-04 | *** |
| 10 | Nucleus cytoplasm ratio | 6.00E-03 | ** |
| 11 | LD number | 3.89E-05 | **** |
| 12 | LD distance | 3.83E-03 | ** |
| 13 | LD relative distance | 1.52E-01 | ns |
| 14 | LD area | 5.22E-06 | **** |
| 15 | LD area ratio | 5.12E-03 | ** |
| 16 | Lipid intensity | 7.62E-01 | ns |
| 17 | Protein intensity | 4.82E-01 | ns |
| 18 | Lipid protein ratio | 7.67E-04 | *** |
| 19 | Lipid intensity within cytoplasm area | 7.31E-01 | ns |
| 20 | Protein intensity within cytoplasm area | 5.76E-01 | ns |

ns: non-significant, *: $p < 0.05$, **: $p < 0.01$, ***: $p < 0.001$, ****: $p < 0.0001$

**Supplementary Table 3. Performance comparisons by PCA-MIL methods.**

| Phenotyping methods | Diagnostic models | SEN (%) | SPE (%) | ACC (%) | PPV (%) | NPV (%) | AUC |
|---|---|---|---|---|---|---|---|
| **PCA-MIL** | LR | 88.46 | 85.19 | 86.25 | 74.19 | 93.88 | 0.907 |
| | SVM | 65.38 | 51.85 | 56.25 | 39.53 | 75.68 | 0.535 |
| | XGBoosting | 92.31 | 87.04 | 88.75 | 77.42 | 95.92 | 0.940 |

**Supplementary Table 4. Summary of concordance between LUCID predictions and clinicopathological characteristics**

| | | LUCID Prediction (%) |
|---|---|---|
| **BCa** | | 92.31 |
| | **Stage** | |
| | Ta | 100.00 |
| | T1 | 88.24 |
| | T2 | 100.00 |
| | T3 | 100.00 |
| | **Grade** | |
| | LG | 90.00 |
| | HG | 93.75 |
| | **Size of tumor** | |
| | ≤1.5 cm | 100.00 |
| | 1.5-3 cm | 75.00 |
| | ≥3 cm | 100.00 |
| | **Number of tumors** | |
| | Single | 100.00 |
| | Multiple | 90.91 |
| **Early-stage of tumor （Ta&T1）** | | 90.91 |
| | Early-stage of tumor-LG | 90.00 |
| | Early-stage of tumor-HG | 91.67 |
| **Non-BCa** | | 87.04 |
| | Normal | 83.33 |
| | Urinary stones | 75.00 |
| | Interstitial cystitis | 66.67 |
| | Urinary incontinence | 75.00 |
| | BPH | 91.89 |
